# Gender/Sex Disparities in the COVID-19 Cascade from Testing to Mortality: An Intersectional Analysis of Swiss Surveillance Data

**DOI:** 10.1101/2024.02.16.24302879

**Authors:** Diane Auderset, Michaël Amiguet, Carole Clair, Valérie Pittet, Julien Riou, Joëlle Schwarz, Yolanda Mueller

## Abstract

**Objectives:** This study investigates gender and sex disparities in COVID-19 epidemiology in the Canton of Vaud, Switzerland, focusing on the interplay with socioeconomic position (SEP) and age.

**Methods:** We analyzed COVID-19 surveillance data from March 2020 to June 2021, using an intersectional approach. Negative binomial regression models assessed disparities between women and men, across SEP quintiles and age groups, in testing, positivity, hospitalizations, ICU admissions, and mortality (Incidence Rate Ratios [IRR], with 95% Confidence Intervals [CI]).

**Results:** Women had higher testing and positivity rates than men, while men experienced more hospitalizations, ICU admissions, and deaths. The higher positivity in women under 50 was mitigated when accounting for their higher testing rates. Within SEP quintiles, gender/sex differences in testing and positivity were not significant. In the lowest quintile, women’s mortality risk was 68% lower (Q1: IRR 0.32, CI 0.20-0.52), with decreasing disparities with increasing SEP quintiles (Q5: IRR 0.66, CI 0.41-1.06).

**Conclusion:** Our findings underscore the complex epidemiological patterns of COVID-19, shaped by the interactions of gender/sex, SEP, and age, highlighting the need for intersectional perspectives in both epidemiological research and public health strategy development.

## INTRODUCTION

The COVID-19 pandemic has had heterogeneous impacts, with certain populations being disproportionately affected. The literature on COVID-19 indicates that socioeconomically disadvantaged groups face higher risks of contracting the virus and experiencing severe outcomes such as hospitalization and mortality (1–5). This risk is linked to socioeconomic determinants, where limited income and education create conditions that elevate exposure risk and susceptibility to infection (1). In many countries, studies have highlighted socioeconomic disparities in the COVID-19 cascade, both nationally (5–8) and regionally (4, 9), indicating that neighborhood-level socioeconomic vulnerability shapes these disparities (10). Additionally, substantial gender differences, particularly in labor and family domains, impact individuals throughout their lifespan, contributing to gendered socioeconomic inequalities that affect health (11).

Globally, men were more likely to develop severe forms of COVID-19, resulting in higher hospitalizations and mortality rates compared to women (12–18). The origins and pathways of these disparities are rooted in a complex interplay of gender-specific social processes and sex-related biological attributes. Gender, as a major social determinant, shapes life experiences and health outcomes through systematic differences in roles, responsibilities, access to power, and opportunities between women and men (19, 20). Gender inequalities in health arise from the intricate interaction of multiple factors, including differential exposure to health risks, health-related behaviors, access to healthcare, and gender biases in healthcare and research (19, 21). Exploring gender and sex reveals nuanced pathways impacting the COVID-19 progression from testing rates to mortality. Gender affects individuals’ health behaviors, access to healthcare, occupational exposures, and adherence to public health measures, thereby potentially affecting COVID-19 exposure, testing rates, positivity, and the burden of disease (15, 22–25). Conversely, sex-related differences, such as immune responses and hormone levels, primarily influence susceptibility, severity, and mortality rates of COVID-19 (13, 26, 27). Studies suggest that hormones like oestrogens and progesterone, typically higher in women, offer protection against viral infections, whereas testosterone, predominant in men, may have the opposite effect (28, 29). Additionally, men often exhibit higher ACE-2 receptor levels, used by SARS-CoV-2 to enter cells, potentially explaining the more severe infection cases (26, 28, 30).

The significant variations in the influence of gender and sex across socioeconomic conditions and over the lifespan underscore the importance of intersectional approaches to deepen our understanding of COVID-19 epidemiology (28, 31). Building on Kimberlé Crenshaw’s work, the concept of intersectionality emerges as a critical framework that clarifies the complex and multifaceted nature of individual experiences, such as their health, shaped by various forms of oppression, including class and gender inequalities (32). An intersectional perspective highlights the intricate ways in which socially constructed categories—rooted in structural power relations— intersect at multiple levels, often independently or simultaneously, to produce nuanced layers of disadvantage (19, 32, 33). It helps to move beyond the consideration of isolated risk factors (34) and focuses on identifying modifiable cause for health inequalities, offering major insights for formulating equitable public health policies and interventions (35).

This study analyzes surveillance data from the canton of Vaud, located in the southwestern part of Switzerland within the French-speaking region. As one of Switzerland’s larger cantons by population and area, it encompasses diverse urban and rural settings, representing approximately one-tenth of the national population. The primary objective is to explore gender and sex disparities in the COVID-19 epidemiology cascade, from testing to mortality, including test positivity, hospitalization, and intensive care unit (ICU) admission. We examined these disparities in the context of key social determinants of health, focusing on neighborhood-based socioeconomic position (SEP) and age, through an intersectional analytical approach. We aim to uncover the complex dynamics underlying these disparities and enhance our understanding of COVID-19’s broader epidemiology. Such knowledge, by considering gender and sex influences, vulnerabilities, and diverse social determinants of health is essential for developing targeted and effective public health strategies to address COVID-19 and guide responses to future pandemics (28).

## METHODS

### Study design and setting

This observational retrospective study analyzed COVID-19 surveillance data from March 2020 to the end of June 2021 of the population residing in the canton of Vaud. The first epidemic wave in Switzerland spanned from February to May 2020, characterized by low testing capacities with RT-PCR tests (4, 5). The tested population primarily included symptomatic individuals, those with known risk factors (e.g., people with comorbidities), and healthcare workers (4). Testing was expanded on June 24, 2020, to include mildly symptomatic individuals and close contacts of infected individuals, with test costs reimbursed (5). Vaccinations began in December 2020, initially for vulnerable groups, and expanded with the opening of vaccination on January 11, 2021 (36, 37). By June 2021, 85% of those aged 75 or older, and 53% of those aged 18 to 49 had received at least one vaccine dose (38).

### Data

Within the Swiss federal state, the Federal Office of Public Health (FOPH) oversees the monitoring of transmissible diseases, including COVID-19, in collaboration with cantonal authorities, through mandatory reporting of infectious diseases (36). Entities authorized to conduct SARS-CoV-2 testing (RT-PCR and rapid antigen tests), such as general practitioners, pharmacies, and testing centers, had to notify each test (negative and positive) to the FOPH. Hospitalizations (lasting at least 24 hours) and ICU admissions due to COVID-19 were reported by hospitals. Probable or confirmed COVID-19-related deaths were reported to cantonal health authorities (see Supplementary Table S1 for case definitions). Population data as of December 31, 2020, were obtained from the cantonal office of statistics (39). The SEP of notified individuals was determined using the Swiss-SEP, an area-based indicator (40, 41). Detailed information on the geocoding procedures is provided in Supplementary Section 2.

The study period spanned 69 weeks from March 2, 2020 (first notified cases in Vaud canton), to June 27, 2021, marking the cessation of the cantonal hospital’s surveillance system. Due to inconsistent negative test reporting prior to May 24, 2020, the dataset for the total number of tests was limited to the period from May 27, 2020, to June 27, 2021, covering a span of 57 weeks. Notifications included the date, test result (positive, negative), date of birth or age, and residential address. Additionally, the gender/sex of each individual was recorded. For hospitalization, death, and PCR test notifications, the gender/sex indicator conformed to the administrative sex categories, restricted to female or male in Switzerland. In contrast, rapid antigenic tests included an “other” option alongside “women” and “men”, acknowledging non-binary gender identities. Consequently, we refer to this variable as “gender/sex” throughout our analysis. Duplicated notifications, records with invalid residential addresses, and those missing age or gender/sex information were excluded. Additionally, notifications with “other” as gender/sex (0.001% of total tests) were excluded. Age was grouped into eight categories of 10-year age bands and 80 and above. For hospitalizations, ICU admissions, and deaths, ages 0 to 59 years were combined due to low number of events in this age range.

### The Swiss socio-economic position (Swiss-SEP)

The Swiss-SEP, an area-based socio-economic position index centered on each residential building and incorporating neighborhood information from the surrounding 50 households, was developed by the Swiss National Cohort (41) (detailed in Supplementary Section 3). The Swiss-SEP index, derived through principal component analysis, aggregates neighborhood-level data from the 2000 census and 2012-2015 annual micro-census. This index utilizes key indicators as proxies for SEP: median rent per square meter (income proxy), proportion of households led by individuals with a primary education or less (education proxy), proportion of households headed by individuals in manual or unskilled jobs (occupation proxy), and the average number of persons per room (crowding proxy). The index scores range from 0 to 100, with higher values indicating higher SEP (40). There are 115 596 SEP neighborhoods within the geographical boundaries of Vaud canton.

Residential coordinates of each notification were matched with the nearest SEP neighborhood, and SEP index values were categorized into quintiles from one (lowest) to five (highest). Non-residential addresses, such as schools or nursing homes, and addresses with only ZIP code information, were assigned the average SEP of their ZIP code area. Regarding total and positive test notifications, 94% and 92% respectively, were successfully geocoded, thus assigned an address-based SEP (See Supplementary Table 4). However, notifications for hospitalization and ICU admission, that contained only ZIP code information, did not receive a SEP assignment due to the method of assigning average SEP scores to ZIP code areas, which tends to centralize distribution around mean values, thereby reducing variability at the extremes of the SEP quintiles. Death notifications that could not be geocoded (39%) were likewise excluded from analyses requiring SEP attribution, due to similar concerns regarding the accuracy of SEP assignment.

### Statistical analysis

The distribution of notifications stratified by gender/sex, across age groups, and SEP quintiles were described. Incidence rates of tests, positive tests, hospitalizations, ICU admissions, and deaths were calculated weekly per 100 000 persons, stratified by gender/sex categories. Cumulative incidence rates over the study period were similarly computed. Negative binomial regression models were used to examine the incidence rate ratios (IRR) with 95% confidence intervals (CIs) between women and men, with interaction terms between sex/gender and age groups and between SEP quintiles. These models, which can handle overdispersion of residuals, included denominators as offsets, with corresponding age and sex structure of the general population as of December 31, 2020, serving as the base for all outcomes. Specifically for positive tests, an additional negative binomial model using the total number of tests as the denominator was formulated to investigate gender/sex-specific test positivity ratios. A similar methodology was applied for ICU admissions, with hospitalizations serving as the offset. Sensitivity analyses were conducted on death notifications, incorporating notifications from institutional locations, followed by a comprehensive analysis of all death notifications, including those not precisely geocoded. Statistical analyses were conducted using R statistical software (42), and negative binomial models estimated using the MASS package (43).

### Data reporting standards

This research aligns with the Sex and Gender Equity in Research (SAGER) guidelines, which advocate for systematic integration of sex and gender considerations into research design, analysis, and reporting (44). Consequently, we will discuss the sex and/or gender-related mechanisms potentially influencing the findings reported within the context of Switzerland. In this paper, the term “gender/sex” is used to acknowledge the complex interplay between these concepts from a theoretical perspective (45). This terminology effectively reflects the varied nature of surveillance data analyzed, where the indicator might represent either administrative sex or gender identity, depending on the notification type. From a methodological standpoint, gender/sex acts as a proxy capturing both gender-related aspects (e.g., behaviors) and sex-related biological factors (e.g., hormonal differences), which may impact the outcomes studied.

## RESULTS

By the end of 2020, Vaud population was 815 300, comprising 412 599 women (50.6%) and 402 701 men (49.4%) (Table 1). From March 2020 to June 2021, a total of 885 925 SARS-CoV-2 tests, 96 963 positive tests, 6 356 hospitalizations, 1 134 ICU admissions and 1 175 deaths (before excluding non-geocoded death notifications) were notified and met eligibility criteria (see Supplementary Figure 5). Although women had more tests and positive results, the majority of hospitalizations, ICU admissions, and deaths occurred among men.

**Table 1.**
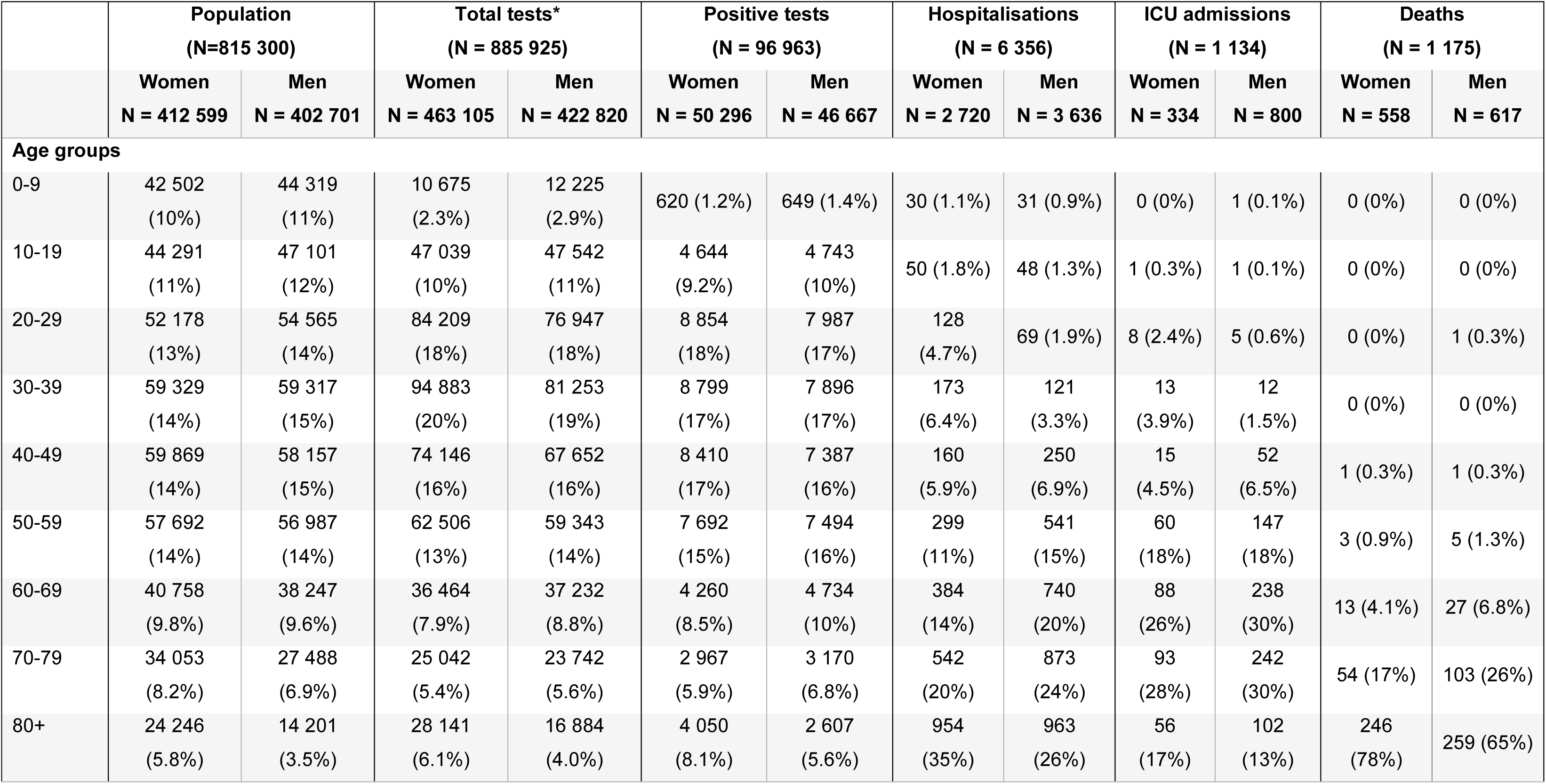
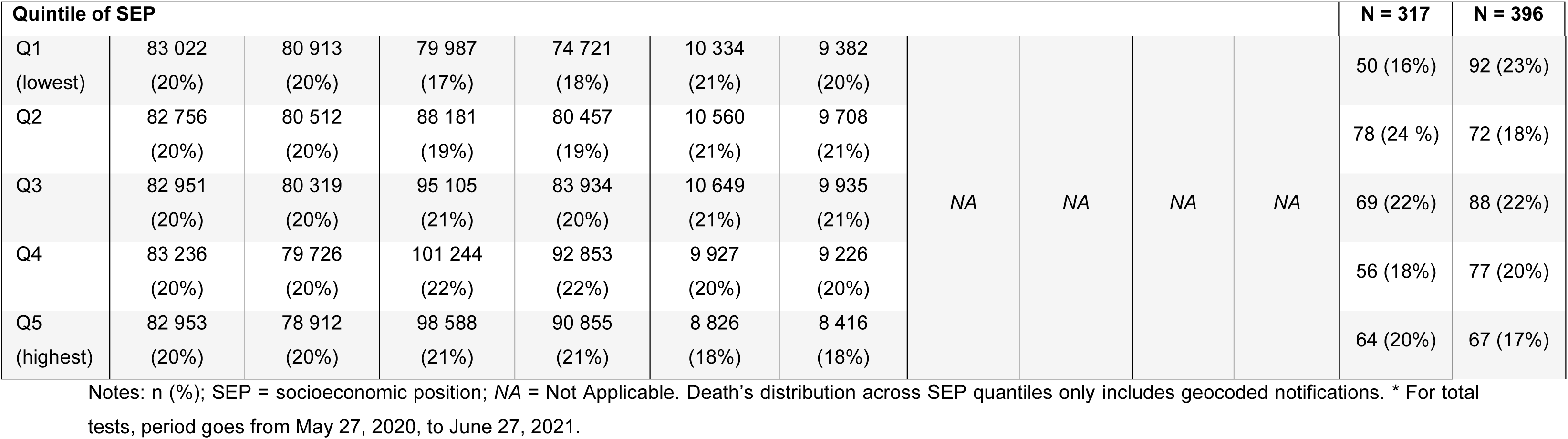
Distribution of age and socioeconomic position (SEP), stratified by gender/sex categories, Canton of Vaud surveillance data, from March 2*, 2020 to June 27, 2021, Switzerland

|  | Population<br>(N=815 300) |  | Total tests*<br>(N = 885 925) |  | Positive tests<br>(N = 96 963) |  | Hospitalisations<br>(N = 6 356) |  | ICU admissions<br>(N = 1 134) |  | Deaths<br>(N = 1 175) |  |
| --- | --- | --- | --- | --- | --- | --- | --- | --- | --- | --- | --- | --- |
|  | Women<br>N = 412 599 | Men<br>N = 402 701 | Women<br>N = 463 105 | Men<br>N = 422 820 | Women<br>N = 50 296 | Men<br>N = 46 667 | Women<br>N = 2 720 | Men<br>N = 3 636 | Women<br>N = 334 | Men<br>N = 800 | Women<br>N = 558 | Men<br>N = 617 |
| **Age groups** |  |  |  |  |  |  |  |  |  |  |  |  |
| 0-9 | 42 502<br>(10%) | 44 319<br>(11%) | 10 675<br>(2.3%) | 12 225<br>(2.9%) | 620 (1.2%) | 649 (1.4%) | 30 (1.1%) | 31 (0.9%) | 0 (0%) | 1 (0.1%) | 0 (0%) | 0 (0%) |
| 10-19 | 44 291<br>(11%) | 47 101<br>(12%) | 47 039<br>(10%) | 47 542<br>(11%) | 4 644<br>(9.2%) | 4 743<br>(10%) | 50 (1.8%) | 48 (1.3%) | 1 (0.3%) | 1 (0.1%) | 0 (0%) | 0 (0%) |
| 20-29 | 52 178<br>(13%) | 54 565<br>(14%) | 84 209<br>(18%) | 76 947<br>(18%) | 8 854<br>(18%) | 7 987<br>(17%) | 128<br>(4.7%) | 69 (1.9%) | 8 (2.4%) | 5 (0.6%) | 0 (0%) | 1 (0.3%) |
| 30-39 | 59 329<br>(14%) | 59 317<br>(15%) | 94 883<br>(20%) | 81 253<br>(19%) | 8 799<br>(17%) | 7 896<br>(17%) | 173<br>(6.4%) | 121<br>(3.3%) | 13<br>(3.9%) | 12<br>(1.5%) | 0 (0%) | 0 (0%) |
| 40-49 | 59 869<br>(14%) | 58 157<br>(15%) | 74 146<br>(16%) | 67 652<br>(16%) | 8 410<br>(17%) | 7 387<br>(16%) | 160<br>(5.9%) | 250<br>(6.9%) | 15<br>(4.5%) | 52<br>(6.5%) | 1 (0.3%) | 1 (0.3%) |
| 50-59 | 57 692<br>(14%) | 56 987<br>(14%) | 62 506<br>(13%) | 59 343<br>(14%) | 7 692<br>(15%) | 7 494<br>(16%) | 299<br>(11%) | 541<br>(15%) | 60<br>(18%) | 147<br>(18%) | 3 (0.9%) | 5 (1.3%) |
| 60-69 | 40 758<br>(9.8%) | 38 247<br>(9.6%) | 36 464<br>(7.9%) | 37 232<br>(8.8%) | 4 260<br>(8.5%) | 4 734<br>(10%) | 384<br>(14%) | 740<br>(20%) | 88<br>(26%) | 238<br>(30%) | 13 (4.1%) | 27 (6.8%) |
| 70-79 | 34 053<br>(8.2%) | 27 488<br>(6.9%) | 25 042<br>(5.4%) | 23 742<br>(5.6%) | 2 967<br>(5.9%) | 3 170<br>(6.8%) | 542<br>(20%) | 873<br>(24%) | 93<br>(28%) | 242<br>(30%) | 54 (17%) | 103 (26%) |
| 80+ | 24 246<br>(5.8%) | 14 201<br>(3.5%) | 28 141<br>(6.1%) | 16 884<br>(4.0%) | 4 050<br>(8.1%) | 2 607<br>(5.6%) | 954<br>(35%) | 963<br>(26%) | 56<br>(17%) | 102<br>(13%) | 246<br>(78%) | 259 (65%) |

| Quintile of SEP |  |  |  |  |  |  |  |  |  |  | N = 317 | N = 396 |
| --- | --- | --- | --- | --- | --- | --- | --- | --- | --- | --- | --- | --- |
| Q1<br>(lowest) | 83 022<br>(20%) | 80 913<br>(20%) | 79 987<br>(17%) | 74 721<br>(18%) | 10 334<br>(21%) | 9 382<br>(20%) | NA | NA | NA | NA | 50 (16%) | 92 (23%) |
| Q2 | 82 756<br>(20%) | 80 512<br>(20%) | 88 181<br>(19%) | 80 457<br>(19%) | 10 560<br>(21%) | 9 708<br>(21%) |  |  |  |  | 78 (24 %) | 72 (18%) |
| Q3 | 82 951<br>(20%) | 80 319<br>(20%) | 95 105<br>(21%) | 83 934<br>(20%) | 10 649<br>(21%) | 9 935<br>(21%) |  |  |  |  | 69 (22%) | 88 (22%) |
| Q4 | 83 236<br>(20%) | 79 726<br>(20%) | 101 244<br>(22%) | 92 853<br>(22%) | 9 927<br>(20%) | 9 226<br>(20%) |  |  |  |  | 56 (18%) | 77 (20%) |
| Q5<br>(highest) | 82 953<br>(20%) | 78 912<br>(20%) | 98 588<br>(21%) | 90 855<br>(21%) | 8 826<br>(18%) | 8 416<br>(18%) |  |  |  |  | 64 (20%) | 67 (17%) |

Notes: n (%); SEP = socioeconomic position; NA = Not Applicable. Death's distribution across SEP quantiles only includes geocoded notifications. \* For total tests, period goes from May 27, 2020, to June 27, 2021.

In the canton of Vaud, 38% of women and 34% of men were aged 50 and above. Among women, this age group accounted for 33% of all tests and 38% of positive tests, but represented 80% of hospitalizations, 89% of ICU admissions, and 99.8% of deaths. Likewise, men aged 50 and older accounted for 32% of tests, 39% of positive tests, 86% of hospitalizations, 91% of ICU admissions, and 99.4% of deaths.

For total tests, 17% of women and 18% of men were in the lowest socioeconomic quintile (Q1), while 21% of tests for both women and men occurred in the highest quintile (Q5). Regarding positive tests, 21% for women and 20% for men were recorded in Q1, with 18% in Q5 for both. In terms of mortality, 18% of men who died were in Q1, and 14% in Q5. Among women, 12% of deaths occurred in Q1 and 15% in Q5.

The weekly incidence of outcomes per 100 000 showed distinct patterns between women and men throughout the study period (Figure 1). Women had higher incidence rate of tests and positive tests compared to men, especially during the second wave of the pandemic. In contrast, men had higher incidence rates of hospitalizations and ICU admissions throughout the study period, though the disparity in mortality rates was less pronounced. During the third wave, while testing rates peaked, both severe outcomes and positivity rates were comparatively lower.

**Figure 1.**
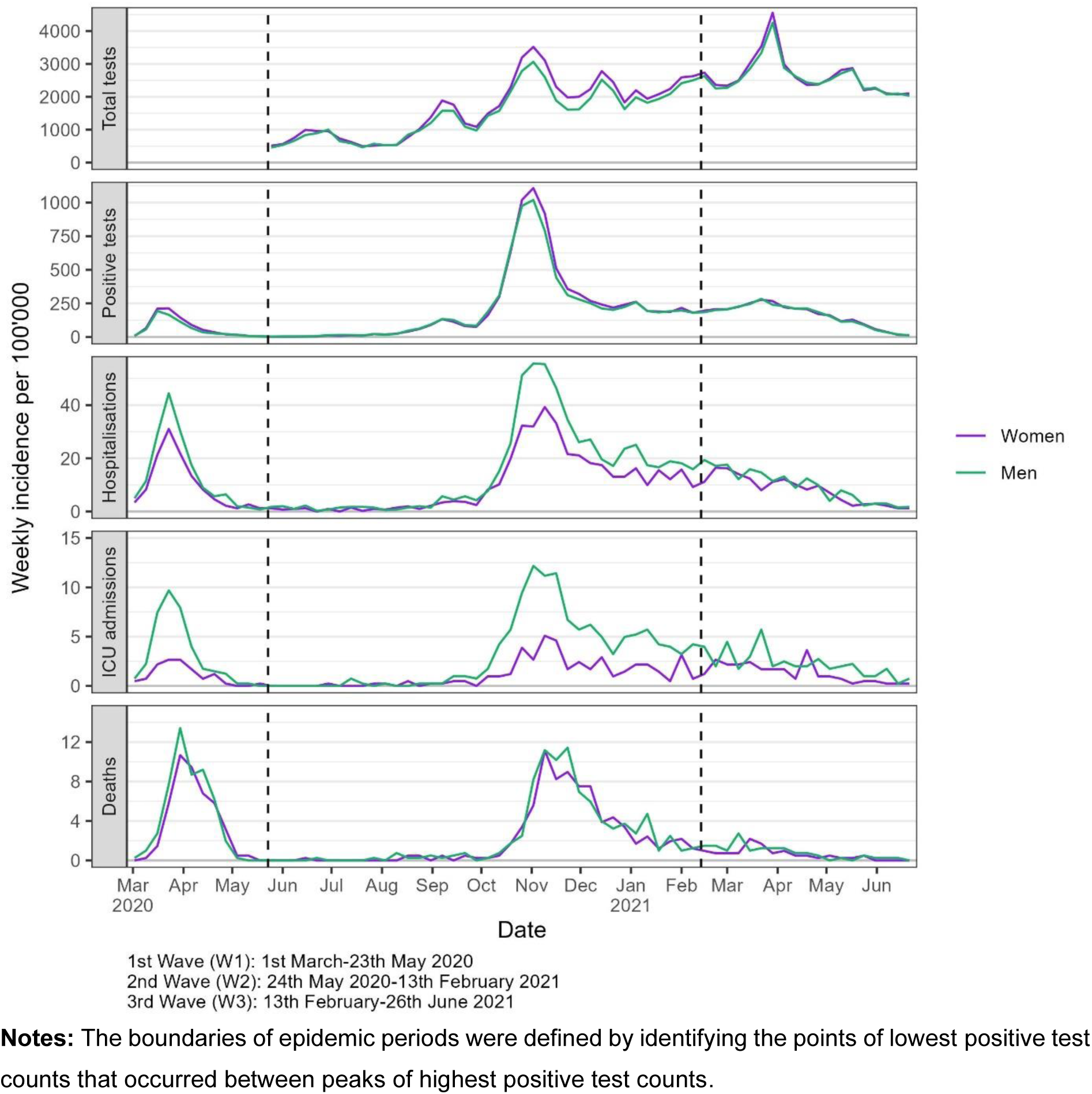
– Weekly incidence of COVID-19 outcomes per 100 000, stratified by gender/sex, Canton of Vaud surveillance data 2020-2021, Switzerland

The cumulative incidences of outcomes across age groups, gender/sex categories, and SEP quintiles revealed distinct patterns (Figure 2). Individuals aged 20 to 39 were the most tested group, whereas children under 10 were the least tested. Testing rates were higher for people aged 80 and above compared to those aged 60-69 and 70-79. Similar patterns emerged across age groups concerning positivity. For severe outcomes, prominent age-related trends were observed, with older age groups experiencing higher incidence rates, though ICU admission were less frequent among those aged 80 and above. Men experienced higher incidence rates of hospitalization, ICU admission, and death than women.

**Figure 2.**
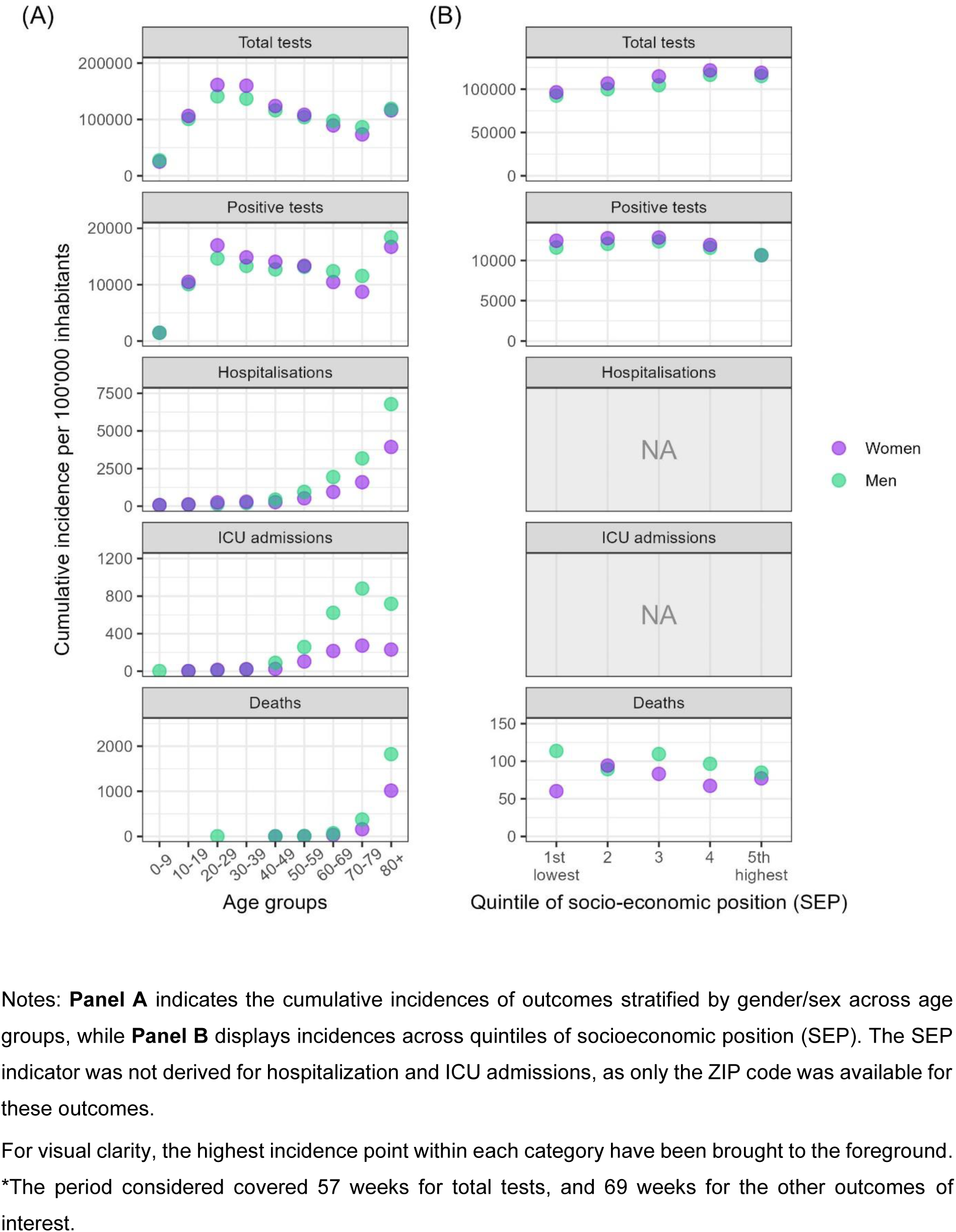
– Cumulative incidence of outcomes between March 2nd, 2020 and June 27, 2021* per 100 000, stratified by gender/sex, across age groups and quintiles of socio-economic position, Canton of Vaud surveillance data 2020-2021, Switzerland

In terms of SEP quintiles (Figure 2, panel B), the cumulative incidence of testing progressively increased from Q1 to Q5. For the cumulative incidence of positive tests, the three lowest SEP quintiles (Q1-Q3) showed similar rates, with lower rates observed in the two highest quintiles (Q4-Q5). A consistent trend was observed, with women having higher cumulative incidence of both tests and positive tests across all quintiles, except in Q5 where the positivity incidence was comparable between women and men. For death notifications, men’s cumulative mortality rate appeared to decrease from Q1 to Q5, whereas for women, the mortality rate was lowest in Q1 and Q4. Men displayed higher mortality rates across all SEP quintiles, except in Q2.

Regression analyses demonstrated distinct testing patterns between women and men across age groups. Notably, women aged 20-29 (IRR 1.14, CI 1.07-1.22) and 30-39 (IRR 1.16, CI 1.09-1.24) displayed a significantly higher likelihood of undergoing testing compared to men in the same age groups (Figure 3, left panel). Conversely, girls under 10 (IRR 0.91, CI 0.85-0.97) and women aged 60-69 (IRR 0.92, CI 0.86-0.98) and 70-79 (IRR 0.85, CI 0.80-0.91) were less likely to get tested compared to their male counterparts. For incidence of positive tests per population, similar gender/sex trends were observed across age groups (Figure 3, center panel). However, these trends were not apparent when adjusting for the initial gender/sex differences in testing, as indicated by the regression results of positive tests per test (Figure 3, right panel). An exception was observed among individuals aged 60 and older, where women were less likely to test positive per test compared to men. Specifically, women had an IRR of 0.92 in the 60-69 age group (CI 0.86-0.98), 0.89 in the 70-79 age group (CI 0.82-0.95), and 0.83 among those aged 80 and older (CI 0.86-1.00), indicating a lower likelihood of a positive result when tested. Moreover, when comparing women and men in similar SEP quintiles (Figure 3, red coefficients), no statistically significant differences in testing and positive testing rates were found, except for women in Q3 who presented a slightly reduced probability of testing positive per test compared to their male counterparts.

**Figure 3.**
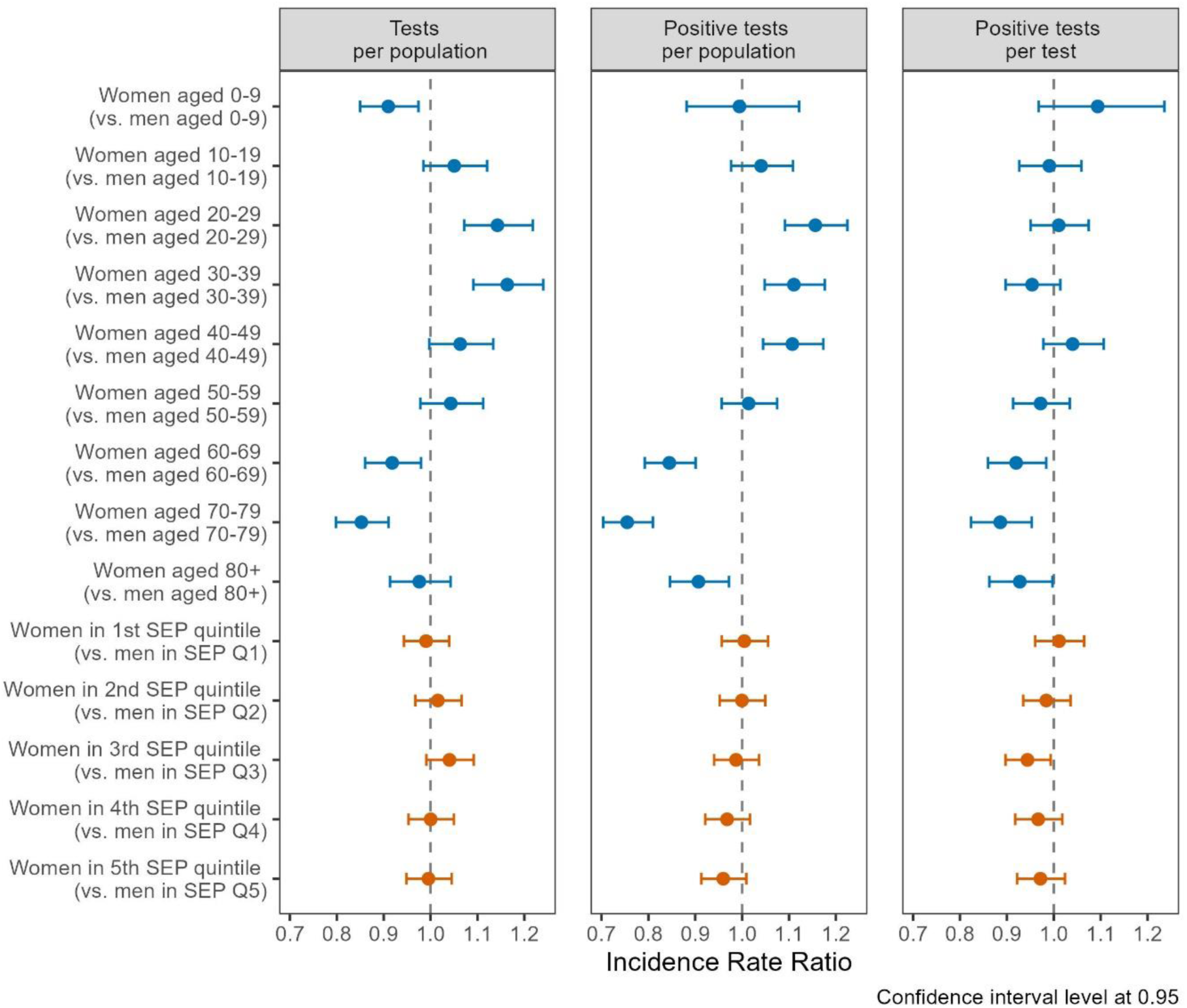
–Incidence rate ratios (IRR) of gender/sex (ref.: men) for number of tests and of positive tests, stratified by age groups (upper part), and quintiles of socio-economic position (SEP, lower part), using general population (left and center panel) and total number of tests (right panel) as denominator, Canton of Vaud surveillance data 2020-2021, Switzerland

In regression models without an interaction term for gender/sex categories (Supplementary Table 6), individuals in Q5 were notably more likely to undergo testing (IRR 1.25, CI 1.19-1.30) compared to those in Q1. Conversely, individuals in Q5 showed a decreased likelihood of testing positive per person (IRR 0.89, CI 0.85-0.95) and testing positive per test (IRR 0.71, CI 0.68-0.74).

Age was the strongest predictor for hospitalisations, ICU admissions and deaths, increasing age being associated with higher likelihoods of these events, as shown by the regression analysis in models without an interaction term by gender/sex (Supplementary Table 6). Analyzing the interaction of gender/sex with age (Figure 4, Panel A), women exhibited lower probabilities of COVID-19 hospitalization than men across all age cohorts, although the differences were not statistically significant. The IRR for women up to 59 was 0.80 (CI 0.52-1.22), 0.49 (CI 0.18-1.36) for those 60-69, 0.50 (CI 0.18-1.38) for the 70-79 age group, and 0.58 (CI 0.21-1.61) for those 80 and above. Statistical significance was achieved for ICU admissions in the under-60 cohort, where women had a 55% decreased risk (IRR 0.45, CI 0.23-0.86). Women consistently showed a lower risk of ICU admissions compared to men when hospitalized (figure 4, Panel B). Women under 60 presented an IRR of 0.59 (CI 0.44-0.78), denoting a 41% lower risk. In the 60-69 age group, the IRR was 0.71 (CI 0.49-1.01), with the risk reduction becoming more pronounced with advancing age. Women aged 70-79 had an IRR of 0.62 (CI 0.43-0.88), while those aged 80 and over had an IRR of 0.56 (CI 0.37-0.85), mirroring the risk reduction observed in the youngest age group.

**Figure 4.**
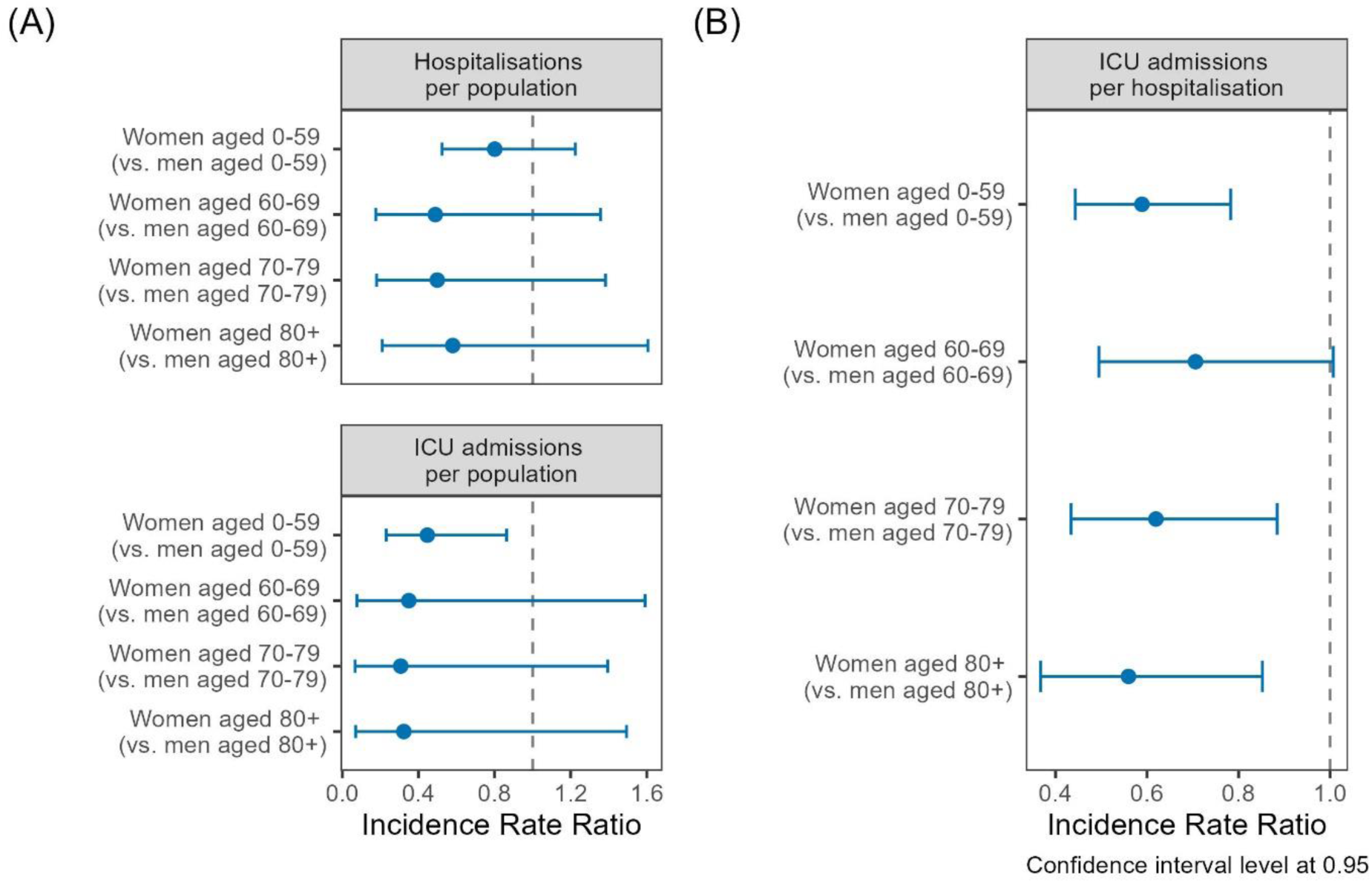
– Incidence rate ratios (IRR) of gender/sex (ref.: men) for hospitalization (upper panel) and ICU admission (lower panel) stratified by age groups, using general population as offset (panel A), and ICU admission per hospitalisations (panel B), Canton of Vaud surveillance data 2020-2021, Switzerland

Regarding mortality, individuals in Q5 had a lower likelihood of death (IRR 0.71, CI 0.54-0.95) compared to those in Q1, as indicated by the regression models without a gender/sex interaction term (Supplementary Table 6). This association between SEP and death persisted in the sensitivity analysis that included nursing home residents and remained robust when extended to include non-precisely geocoded death notifications (Supplementary material S7). Lower mortality rates among women were noted across all age groups (figure 5). Women demonstrated a reduced mortality risk compared to men of 55% at ages 60-69 (IRR 0.45, CI 0.23-0.88), 58% at ages 70-79 (IRR 0.42, CI 0.30-0.59), and 45% for those aged 80 and above (IRR 0.55, CI 0.46-0.66). Regarding SEP, the gender/sex disparities in mortality were more pronounced in Q1, with women having a 68% reduction in mortality risk (IRR 0.32, CI 0.20-0.52); and these disparities were not statistically significant in Q2 and Q5. Exploring the combined effects of gender/sex, age, and SEP on mortality among older groups (70-79 and 80+), a triple interaction term was employed (Supplementary Figure S8). Our findings indicate a reduction in gender/sex mortality disparities with increasing SEP, as the IRR tends toward unity from the lowest to highest quintiles.

**Figure 5.**
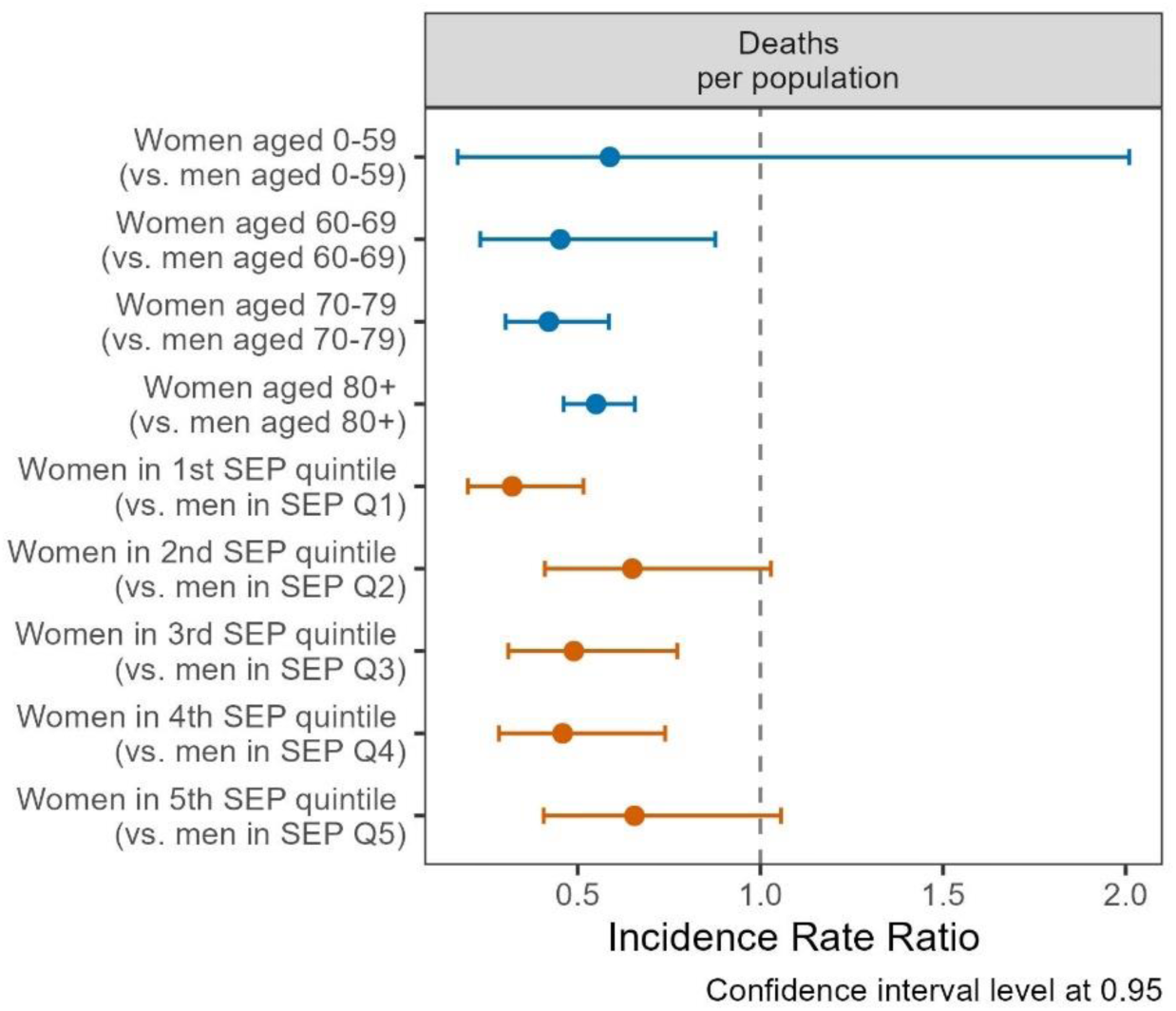
– Incidence rate ratios (IRR) of gender/sex (ref.: men) for death, by age groups (blue coefficients), and quintiles of socio-economic position (SEP, red coefficients), using general population as offset, Canton of Vaud surveillance data 2020-2021, Switzerland

## DISCUSSION

In the resident population of the Canton of Vaud, Switzerland, women contributed to a higher number of COVID-19 tests and positive tests than men, whereas more hospitalizations, ICU admissions, and deaths occurred among men. This finding underscores a pronounced gender/sex disparity in the pandemic’s health impact, highlighting the need to explore underlying causes, such as potential biological differences, gender-specific behavioral patterns, and occupational exposures.

Individuals residing in the highest SEP neighborhoods underwent more COVID-19 testing than those in the lowest SEP areas, accompanied by a lower likelihood of testing positive and a reduced risk of mortality. These observations suggest significant socio-economic influences on health-related behaviors and resource accessibility and utilization. Notably, our intersectional analysis revealed that these disparities in testing and positivity rates are consistent across women and men within similar SEP quintiles. Moreover, the gender/sex disparities in mortality across SEP quintiles highlight the intricate interplay between socioeconomic factors and gender/sex, reinforcing the value of an intersectional approach in uncovering nuanced aspects of COVID-19 epidemiology.

Moreover, age-related variations in SARS-CoV-2 testing rates between women and men were evident in our data. Women aged 20-29 and 30-39 had higher testing rates than men in corresponding age groups, whereas this trend reversed in in the younger (<20) and older age groups (60-69 and 70-79), echoing trends observed in other European countries (46). These variations illustrate a complex relationship between age, gender/sex, and health-seeking behavior, calling for further investigations.

Age is a key factor in understanding COVID-19 gender/sex disparities. The influence of gender norms on health outcomes varies across the life course (19, 28, 31), and the disparities in testing rates between men and women across different age groups likely reflect the evolving societal roles and responsibilities (47). In the 20 to 40 age range, where gender differences in testing were most pronounced, marked distinctions in family and employment domains are generally observed. Women are more likely to work in essential service sectors involving close contacts and limited telecommuting options, such as in service and healthcare jobs (47–50), which may account for their higher testing rates. Yet, this potential increased exposure did not translate into higher positivity rates when accounting for initial differences in testing, possibly attributable to greater adherence to health recommendations and protective measures among women compared to men (23, 26, 29, 49, 51). Additionally, women in this age group often bear a disproportionate burden of unpaid care responsibilities, likely influencing their decisions regarding COVID-19 testing (29, 52). The observed higher testing rates in men aged 60 and above may be attributed to the preferential ascertainment of severe cases (53), where individuals who are more likely, or perceived as more likely–due to early widely reported higher mortality rates in men–, to suffer from severe forms of infection tend to get tested more frequently. Although the ratios of positive tests were generally similar, women aged 60 and above were significantly less likely to test positive per test conducted compared to their male counterparts, highlighting possible differences in exposure.

Our study corroborates the well-established correlation between age and severe COVID-19 outcomes, with older age associated with an increased risk of hospitalization, ICU admissions, and mortality. Additionally, our data confirm that men face a higher risk of severe outcomes compared to women, aligning with previous research (54–56). Notably, women under the age of 60 and those aged 70 and above had a reduced risk of ICU admissions when hospitalized, suggesting possible variations in immune system responses, prevalence of comorbidities, health-seeking behaviors, or differences in treatment approaches between women and men.

Our findings are consistent with existing literature on the link between SEP and COVID-19 outcomes (1–5), highlighting the increased vulnerability of individuals residing in low SEP neighborhoods. This vulnerability stems from a combination of factors, including limited access to healthcare, higher exposure risks due to living and working conditions, occupational hazards, and lifestyle habits, coupled with higher comorbidities rates (57). Previous studies show that those in lower SEP areas experienced lower testing rates–particularly pronounced in the pandemic’s early stages–and faced elevated rates of case incidence, hospitalizations, and mortality, a trend reported globally (1–5, 9, 58–60).

As extensively documented in the literature and corroborated by our findings, men experienced higher mortality rates related to COVID-19 compared to women (12, 13, 15–17, 23, 29, 61). Our results outlined gender/sex disparities in mortality across SEP quintiles, particularly marked in the lowest SEP neighborhoods. This suggests that men from socioeconomically deprived backgrounds may encounter cumulative disadvantages that amplify their health vulnerabilities throughout their lives (62). Biological sex-related factors are thought to play a significant role in men’s increased vulnerability to COVID-19, possibly mediated through hormonal and immune response (29, 50). Mortality is also influenced by gendered practices and societal norms such as expressions of masculinity, which intersect with various social determinants of health (50). These include working in hazardous industries, engagement in risky health behaviors, and maintaining lifestyles that lead to higher prevalences of chronic diseases (26, 28, 29, 63, 64), which are more common in socioeconomically disadvantaged populations (58). Beyond age, pre-existing medical conditions such as obesity, diabetes, or chronic respiratory and cardiovascular diseases have been described as major contributors to COVID-19 severity (65). Comparatively riskier health behaviors among men, such as smoking, excessive alcohol consumption, and unhealthy diets, combined with societal norms that valorize toughness and discourage timely medical care, are thought to contribute to their health vulnerabilities (16, 19, 26). Our findings align with existing studies using an intersectional framework, which are notably concentrated in the US and often emphasize ethnic/racial disparities alongside gender/sex, though not always directly correlating these with socioeconomic class distinctions. Notably, a US study found that Black women experienced higher mortality rates than White men, while Black men had the highest mortality rates (66). Another study exploring the combined effects of gender, SEP, and race/ethnicity revealed that, compared to high SEP White women, low SEP White men experienced a mortality rate 7.4 times higher, and 1.5 times higher compared to women in low SEP. Meanwhile, low SEP Hispanic men faced the most significant disparity, with mortality rates 27 times higher than those of high SEP White women (67).

Our study’s strengths include the use of a neighborhood-based SEP indicator to capture potential individual and local-level effects on outcomes, and the minimization of selection bias by using comprehensive surveillance data for the entire Canton of Vaud population. Although Vaud was heavily impacted during the early stages of the pandemic, its diverse rural and urban population profiles provide valuable insights that may reflect broader trends in Switzerland, despite some regional variations. However, our analysis is limited by the absence of data on key individual-level factors such as migration background status and ethnicity. Incorporating these factors could greatly enrich our understanding, especially given that approximately one-third of Vaud’s population in 2020 held non-Swiss nationality (39), and ethnic minorities faced higher exposure and vulnerability to COVID-19 (60, 66, 68). Furthermore, disentangling the sources of disparities between gender and sex is methodologically challenging when using administrative sex to investigate women’s and men’s health outcomes. Nevertheless, our intersectional approach facilitated the development of hypotheses about gendered mechanisms, which extend beyond the traditional biological interpretations common in biomedical research. Another limitation concerns hospitalization and ICU admission data, which may be subject to underreporting due to challenges associated with identifying primary causes of hospitalization, especially among older adults with comorbidities (69). Moreover, deaths occurring outside clinical settings frequently remain untested, complicating their classification as COVID-19 related (69, 70). Additionally, the reliance on residence location for the Swiss-SEP indicator may not accurately reflect an individual’s lifelong SEP, a common challenge with area-based indicators (71).

## Conclusion

Our study within the Canton of Vaud highlighted the significant interplay of gender/sex, age, and SEP in shaping the epidemiology COVID-19. The intersectional analyses have revealed nuanced disparities, notably the increased risk of mortality in men, particularly those from lower SEP neighborhoods. While no substantial gender/sex differences in testing outcomes were observed across SEP quintiles, important age-related variation emerged, with young adult women experiencing higher testing rates. These findings underscore the importance of adopting intersectional approaches in both epidemiological research and public health strategy development. Such approaches are necessary for developing more effective and equitable health responses.

## Data Availability

All data presented in this study are available upon reasonable request to the authors, subject to any applicable privacy or ethical restrictions.

## References

1. Laajaj R, Webb D, Aristizabal D, Behrentz E, Bernal R, Buitrago G, et al. Understanding how socioeconomic inequalities drive inequalities in COVID-19 infections. Scientific Reports. (2022);12(1):8269.

2. Benita F, Rebollar-Ruelas L, Gaytán-Alfaro ED. What have we learned about socioeconomic inequalities in the spread of COVID-19? A systematic review. Sustainable Cities and Society. (2022);86:104158.

3. Vandentorren S, Smaïli S, Chatignoux E, Maurel M, Alleaume C, Neufcourt L, et al. The effect of social deprivation on the dynamic of SARS-CoV-2 infection in France: a population-based analysis. The Lancet public health. (2022);7(3):e240–e9.

4. Mongin D, Cullati S, Kelly-Irving M, Rosselet M, Regard S, Courvoisier DS. Neighbourhood socio-economic vulnerability and access to COVID-19 healthcare during the first two waves of the pandemic in Geneva, Switzerland: A gender perspective. EClinicalMedicine. (2022);46:101352.

5. Riou J, Panczak R, Althaus CL, Junker C, Perisa D, Schneider K, et al. Socioeconomic position and the COVID-19 care cascade from testing to mortality in Switzerland: a population-based analysis. The Lancet Public Health. (2021);6(9):e683–e91.

6. Khazanchi R, Beiter ER, Gondi S, Beckman AL, Bilinski A, Ganguli I. County-Level Association of Social Vulnerability with COVID-19 Cases and Deaths in the USA. J Gen Intern Med. (2020);35(9):2784–7.

7. Berchet C, Bijlholt J, Ando M. Socio-economic and ethnic health inequalities in COVID-19 outcomes across OECD countries. (2023).

8. Jassat W, Ozougwu L, Munshi S, Mudara C, Vika C, Arendse T, et al. The intersection of age, sex, race and socio-economic status in COVID-19 hospital admissions and deaths in South Africa (with corrigendum). South African Journal of Science. (2022);118(5/6).

9. Khan MM, Odoi A, Odoi EW. Geographic disparities in COVID-19 testing and outcomes in Florida. BMC Public Health. (2023);23(1):79.

10. Tudor Hart J. THE INVERSE CARE LAW. The Lancet. (1971);297(7696):405–12.

11. Corna LM. A life course perspective on socioeconomic inequalities in health: A critical review of conceptual frameworks. Advances in Life Course Research. (2013);18(2):150–9.

12. Thompson K, Vassallo A, Finfer S, Woodward M. Renewed rationale for sex-and gender-disaggregated research: A COVID-19 commentary review. Women’s Health. (2022);18:17455065221076738.

13. Gebhard C, Regitz-Zagrosek V, Neuhauser HK, Morgan R, Klein SL. Impact of sex and gender on COVID-19 outcomes in Europe. Biology of Sex Differences. (2020);11(1):29.

14. Scully EP, Schumock G, Fu M, Massaccesi G, Muschelli J, Betz J, et al. Sex and Gender Differences in Testing, Hospital Admission, Clinical Presentation, and Drivers of Severe Outcomes From COVID-19. Open Forum Infectious Diseases. (2021);8(9).

15. Peckham H, de Gruijter NM, Raine C, Radziszewska A, Ciurtin C, Wedderburn LR, et al. Male sex identified by global COVID-19 meta-analysis as a risk factor for death and ITU admission. Nature communications. (2020);11(1):1–10.

16. Bambra C, Albani V, Franklin P. COVID-19 and the gender health paradox. Scandinavian Journal of Public Health. (2021);49(1):17–26.

17. Sharma G, Volgman AS, Michos ED. Sex Differences in Mortality From COVID-19 Pandemic. JACC: Case Reports. (2020);2(9):1407–10.

18. Bauer GR. Sex and Gender Multidimensionality in Epidemiologic Research. Am J Epidemiol. (2023);192(1):122–32.

19. Heise L, Greene ME, Opper N, Stavropoulou M, Harper C, Nascimento M, et al. Gender inequality and restrictive gender norms: framing the challenges to health. The Lancet. (2019);393(10189):2440–54.

20. Kaufman MR, Eschliman EL, Karver TS. Differentiating sex and gender in health research to achieve gender equity. Bull World Health Organ. (2023);101(10):666–71.

21. Shannon G, Jansen M, Williams K, Cáceres C, Motta A, Odhiambo A, et al. Gender equality in science, medicine, and global health: where are we at and why does it matter? Lancet. (2019);393(10171):560–9.

22. Laster Pirtle WN, Wright T. Structural Gendered Racism Revealed in Pandemic Times: Intersectional Approaches to Understanding Race and Gender Health Inequities in COVID-19. Gender & Society. (2021);35(2):168–79.

23. Danielsen AC, Lee KM, Boulicault M, Rushovich T, Gompers A, Tarrant A, et al. Sex disparities in COVID-19 outcomes in the United States: Quantifying and contextualizing variation. Soc Sci Med. (2022);294:114716.

24. Clark C, Davila A, Regis M, Kraus S. Predictors of COVID-19 voluntary compliance behaviors: An international investigation. Global Transitions. (2020);2:76–82.

25. Wenham C, Smith J, Morgan R. COVID-19: the gendered impacts of the outbreak. The lancet. (2020);395(10227):846–8.

26. Ya’qoub L, Elgendy IY, Pepine CJ. Sex and gender differences in COVID-19: More to be learned! American Heart Journal Plus: Cardiology Research and Practice. (2021);3:100011.

27. Groban L, Wang H, Sun X, Ahmad S, Ferrario CM. Is Sex a Determinant of COVID-19 Infection? Truth or Myth? Current Hypertension Reports. (2020);22(9):62.

28. Taslem Mourosi J, Anwar S, Hosen MJ. The sex and gender dimensions of COVID-19: A narrative review of the potential underlying factors. Infection, Genetics and Evolution. (2022);103:105338.

29. Díaz-Rodríguez N, Binkytė R, Bakkali W, Bookseller S, Tubaro P, Bacevičius A, et al. Gender and sex bias in COVID-19 epidemiological data through the lens of causality. Information Processing & Management. (2023);60(3):103276.

30. Youth Repeatedly Hospitalized for DKA: Proof of Concept for Novel Interventions in Children’s Healthcare (NICH) | Diabetes Care.

31. Notarte KI, de Oliveira MHS, Peligro PJ, Velasco JV, Macaranas I, Ver AT, et al. Age, Sex and Previous Comorbidities as Risk Factors Not Associated with SARS-CoV-2 Infection for Long COVID-19: A Systematic Review and Meta-Analysis. Journal of Clinical Medicine. (2022);11(24):7314.

32. Collins PH. Intersectionality’s Definitional Dilemmas. Annual Review of Sociology. (2015);41(1):1–20.

33. Olanlesi-Aliu A, Tulli M, Kemei J, Bonifacio G, Reif LC, Cardo V, et al. A scoping review on the operationalization of intersectional health research methods in studies related to the COVID-19 pandemic. International Journal of Qualitative Studies on Health and Well-being. (2024);19(1):2302305.

34. Molenaar J. Using an intersectional lens to understand the unequal impact of the COVID-19 pandemic. Bi-monthly report 5 September 2021. (2021).

35. Bauer GR. Incorporating intersectionality theory into population health research methodology: Challenges and the potential to advance health equity. Social Science & Medicine. (2014);110:10–7.

36. Giachino M, Valera CBG, Rodriguez Velásquez S, Dohrendorf-Wyss MA, Rozanova L, Flahault A. Understanding the Dynamics of the COVID-19 Pandemic: A Real-Time Analysis of Switzerland’s First Wave. International Journal of Environmental Research and Public Health. (2020);17(23):8825.

37. Office fédéral de la santé publique, Commission fédérale pour les vaccinations. COVID-19 : stratégie de vaccination (2022) 29.11.2022. Available from: https://www.bag.admin.ch/dam/bag/fr/dokumente/mt/k-und-i/aktuelle-ausbrueche-pandemien/2019-nCoV/impfstrategie-bag-ekif.pdf.download.pdf/strategie-de-vaccination-covid-19-ofsp-ekif.pdf.

38. EPICOVID, Direction Générale de la santé (DGS), Office du médecin cantonal. COVID-19: Point épidémiologique – Situation au 28 juin 2021. Available from: https://infosan.vd.ch/fileadmin/2-PUBLICATIONS/SANTE_POPULATION/SSP_20210628_COVID_Bulletin_hebdomadaire_epidemio.pdf.

39. Statistique Vaud (2020). Population résidante permanente par âge exact_ sexe et origine_ Vaud_ 2017-2020 [Available from: https://www.vd.ch/themes/etat-droit-finances/statistique/statistiques-par-domaine/01-population/etat-et-structure-de-la-population.

40. Panczak R, Berlin C, Voorpostel M, Zwahlen M, Egger M. The Swiss neighbourhood index of socioeconomic position: update and re-validation. Swiss Medical Weekly. (2023);153(1):40028.

41. Panczak R, Galobardes B, Voorpostel M, Spoerri A, Zwahlen M, Egger M. A Swiss neighbourhood index of socioeconomic position: development and association with mortality. J Epidemiol Community Health. (2012);66(12):1129–36.

42. R Core Team. R: A language and environment for statistical computing. In: Computing RFfS, editor. Vienna, Austria(2022).

43. Ripley B, Venables B. Modern Applied Statistics with S. Fourth ed. New York: Springer; (2022).

44. Heidari S, Babor TF, De Castro P, Tort S, Curno M. Sex and Gender Equity in Research: rationale for the SAGER guidelines and recommended use. Res Integr Peer Rev. (2016);1(1):2.

45. Fausto-Sterling A. Gender/Sex, Sexual Orientation, and Identity Are in the Body: How Did They Get There? J Sex Res. (2019);56(4-5):529–55.

46. Sobotka T, Brzozowska Z, Muttarak R, Zeman K, di Lego V. Age, gender and COVID-19 infections. medRxiv. (2020):2020.05.24.20111765.

47. Yavorsky JE, Qian Y, Sargent AC. The gendered pandemic: The implications of COVID-19 for work and family. Sociol Compass. (2021);15(6):e12881.

48. Kley S, Reimer T. Exploring the Gender Gap in Teleworking from Home. The Roles of Worker’s Characteristics, Occupational Positions and Gender Equality in Europe. Social Indicators Research. (2023);168(1):185–206.

49. Sant Fruchtman C, Fischer FB, Monzón Llamas L, Tavakkoli M, Cobos Muñoz D, Antillon M. Did COVID-19 Policies Have the Same Effect on COVID-19 Incidence Among Women and Men? Evidence From Spain and Switzerland. Int J Public Health. (2022);67:1604994.

50. Morgan R, Baker P, Griffith DM, Klein SL, Logie CH, Mwiine AA, et al. Beyond a zero-sum game: how does the impact of COVID-19 vary by gender? Frontiers in Sociology. (2021):126.

51. Galasso V, Pons V, Profeta P, Becher M, Brouard S, Foucault M. Gender differences in COVID-19 attitudes and behavior: Panel evidence from eight countries. Proceedings of the National Academy of Sciences. (2020);117(44):27285–91.

52. Bühler N, Pralong M, Rawlinson C, Gonseth S, D’Acremont V, Bochud M, et al. Caring during COVID-19: Reconfigurations of gender and family relations during the pandemic in Switzerland. Frontiers in Sociology. (2021);6:737619.

53. Lipsitch M, Donnelly CA, Fraser C, Blake IM, Cori A, Dorigatti I, et al. Potential Biases in Estimating Absolute and Relative Case-Fatality Risks during Outbreaks. PLoS Negl Trop Dis. (2015);9(7):e0003846.

54. Hawkes S, Pantazis A, Purdie A, Gautam A, Kiwuwa-Muyingo S, Buse K, et al. Sex-disaggregated data matters: tracking the impact of COVID-19 on the health of women and men. Econ Polit (Bologna). (2022);39(1):55–73.

55. Pivonello R, Auriemma RS, Pivonello C, Isidori AM, Corona G, Colao A, et al. Sex Disparities in COVID-19 Severity and Outcome: Are Men Weaker or Women Stronger? Neuroendocrinology. (2021);111(11):1066–85.

56. Scully EP, Schumock G, Fu M, Massaccesi G, Muschelli J, Betz J, et al., editors. Sex and gender differences in testing, hospital admission, clinical presentation, and drivers of severe outcomes from COVID-19. Open forum infectious diseases; 2021: Oxford University Press US; (2021)Published.

57. Acebillo-Baqué M, Maestripieri L. Intersectionality Theory and Its Application in the COVID-19 Pandemics. In: Liamputtong P, editor. Handbook of Social Sciences and Global Public Health. Cham: Springer International Publishing; (2023). p. 1189–211.

58. McGowan VJ, Bambra C. COVID-19 mortality and deprivation: pandemic, syndemic, and endemic health inequalities. The Lancet Public Health. (2022);7(11):e966–e75.

59. Bambra C. Pandemic inequalities: emerging infectious diseases and health equity. International Journal for Equity in Health. (2022);21(1):6.

60. Khanijahani A, Iezadi S, Gholipour K, Azami-Aghdash S, Naghibi D. A systematic review of racial/ethnic and socioeconomic disparities in COVID-19. International Journal for Equity in Health. (2021);20(1):248.

61. Kim H, Fox AM, Kim Y, Kim R, Bae G, Kang M. Is the male disadvantage real? Cross-national variations in sex gaps in COVID-19 incidence and mortality. Glob Public Health. (2021);16(12):1793–803.

62. Turrell G, Lynch JW, Leite C, Raghunathan T, Kaplan GA. Socioeconomic disadvantage in childhood and across the life course and all-cause mortality and physical function in adulthood: evidence from the Alameda County Study. Journal of Epidemiology and Community Health. (2007);61(8):723–30.

63. Anderegg N, Panczak R, Egger M, Low N, Riou J. Survival among people hospitalized with COVID-19 in Switzerland: a nationwide population-based analysis. BMC Med. (2022);20(1):164.

64. Nikoloski Z, Alqunaibet AM, Alfawaz RA, Almudarra SS, Herbst CH, El-Saharty S, et al. Covid-19 and non-communicable diseases: evidence from a systematic literature review. BMC Public Health. (2021);21(1):1068.

65. Agodi A, Maugeri A, Favara G, Magnano San Lio R, Puglisi M, Sinatra D, et al. Gender differences in comorbidities of patients with COVID-19: An Italian local register-based analysis. Heliyon. (2023);9(7):e18109.

66. Rushovich T, Boulicault M, Chen JT, Danielsen AC, Tarrant A, Richardson SS, et al. Sex Disparities in COVID-19 Mortality Vary Across US Racial Groups. Journal of General Internal Medicine. (2021);36(6):1696–701.

67. Pathak EB, Menard JM, Garcia RB, Salemi JL. Joint Effects of Socioeconomic Position, Race/Ethnicity, and Gender on COVID-19 Mortality among Working-Age Adults in the United States. Int J Environ Res Public Health. (2022);19(9).

68. Irizar P, Pan D, Kapadia D, Bécares L, Sze S, Taylor H, et al. Ethnic inequalities in COVID-19 infection, hospitalisation, intensive care admission, and death: a global systematic review and meta-analysis of over 200 million study participants. eClinicalMedicine. (2023);57.

69. Akter S. The gender gap in COVID-19 mortality in the United States. Feminist Economics. (2021);27(1-2):30–47.

70. Riou J, Hauser A, Fesser A, Althaus CL, Egger M, Konstantinoudis G. Direct and indirect effects of the COVID-19 pandemic on mortality in Switzerland. Nature Communications. (2023);14(1):90.

71. Bryere J, Pornet C, Copin N, Launay L, Gusto G, Grosclaude P, et al. Assessment of the ecological bias of seven aggregate social deprivation indices. BMC Public Health. (2017);17(1):86.

