## Supplementary material for "Gender/Sex Disparities in the COVID-19 Cascade from Testing to Mortality: An Intersectional Analysis of Swiss Surveillance Data"

### Supplementary Table 1: Case definitions and notification process of outcomes, cantonal surveillance data of canton of Vaud, 2020-2021, Switzerland

| Outcome | Case definition |
|---|---|
| **Total tests** | Tests reported to the Federal Office of Public Health (FOPH) as per FOPH's declaration criteria, as defined in the Ordinance of the Federal Department of Inner Affairs on declarations of observations related to transmissible diseases in humans (Ordonnance sur la déclaration d'observations en rapport avec les maladies transmissibles de l'homme 818.101.126) and attributed to the canton of Vaud. |
| **Positive tests** | Laboratory-confirmed tests (positive RT-PCR test or rapid antigen test) among total tests notified to the FOPH. |
| **Hospitalisations and Intensive Care Unit (ICU) admissions** | Direct reporting from hospitals and clinics to the Office du Médecin Cantonal (OMC) through the EPICOVID cantonal surveillance system. Included records cover hospital stays in a healthcare facility of the canton of Vaud or in an intercantonal facility. Up to April 15th, 2020, any stay of a patient with a positive SARS-CoV-2 RT-PCR test was included. From April 15th, 2020, inclusion criteria were refined to any stay of a patient with at least one positive RT-PCR test for SARS-CoV-2 and a viral load > 1000 copies during the hospital stay. Admission dates correspond to the entire stay, independent of the date of the viral load test or end of isolation. Each new stay is re-analyzed and included according to these definitions.<br><br>Excluded records:<br>- Hospital stays below 24hrs<br>- Stays in Centre de Traitement et Réadaptation (CTR) or in mental health facility<br>- For patients admitted to the Centre Hospitalier Universitaire Vaudois (CHUV – Lausanne University Hospital), those who opted out of the use of their data for research purposes via the "consentement general" were excluded (N=510)<br>Note: Inclusion criteria vary based on the possibility of automated data extraction from hospitals. Criteria presented above correspond to those of CHUV. For other hospitals, inclusion criteria align with mandatory reporting criteria formulated by the Swiss Federal Office of Public Health (OFSP) as follows (as of 18.05.2020):<br><br>Patient hospitalized for more than 24 hours who presents:<br>• Clinical criteria compatible with COVID-19 and a positive SARS-CoV-2 PCR test, or<br>• Clinical criteria and CT-Scan imaging compatible with COVID-19 and a negative SARS-CoV-2 PCR test without any other known aetiology, or<br>• Clinical and epidemiological criteria and a negative SARS-CoV-2 test without any other known aetiology. |
| **Deaths** | Deaths among probable or confirmed COVID-19 cases reported to the cantonal authorities. |

### Supplementary Section 2: Geocoding procedures and socio-economic position (SEP) attribution

Residential addresses were geocoded using publicly available data of the Federal Register of Buildings and Dwellings (under ORegBL ; RS 431.841) obtained from the Federal Office of Statistics (1). This register contains all residential addresses of Switzerland, at the cantonal level, with corresponding geographic coordinates based on the Swiss LV95 reference system. The process of geocoding and SEP attribution is detailed in figure 2B, next page. Geocoding was performed using both perfect and fuzzy matching methods. Fuzzy matching involves comparing and matching text strings based on their similarity, taking into account variations in word order, spelling, and other factors. The *fuzzyjoin* package in R (2) was used, and distance between sequences of addresses calculated with the *Jaro-Winkler similarity method*, which provides a score between 0 (perfect similarity) and 1 (total dissimilarity). To derive the socio-economic position (SEP) score, ranging from 0 to 100, the residential coordinates in the notifications were matched with the nearest SEP neighborhood using the *SF* package of R (3).

The spatial distribution of the Swiss-SEP index within the Vaud population was estimated to define the SEP cut-off quintiles and to establish the population dataset employed as the denominator for our analyses. This estimation employed Statpop hectometric population data aggregated by age groups (5 years age-range) and sex/gender categories (women, men) (4). All geographic information system processes were performed using the software QGIS (5).

To determine the distribution of SEP within the canton of Vaud's population, hectare-level aggregated data was utilized. For each hectare (depicted as purple squares in the illustration below), SEP values were retrieved or calculated – using the mean of SEP values when multiple SEP neighborhoods fell within a hectare – depicted as yellow dots in Figure 2A. These SEP values were then attributed to the respective population residing within each hectare. SEP values of the population were categorized into five groups using quintiles as cut-offs, ranging from one (lowest) to five (highest). These quintile cut-offs were then used to classify SEP scores of data notifications and assign them to the corresponding quintiles.

**Supplementary Figure 2A: Illustration of hectometric population data (purple squares) and SEP neighborhoods (yellow dots, among the 115'596 SEP neighborhoods in Vaud) on QGIS**

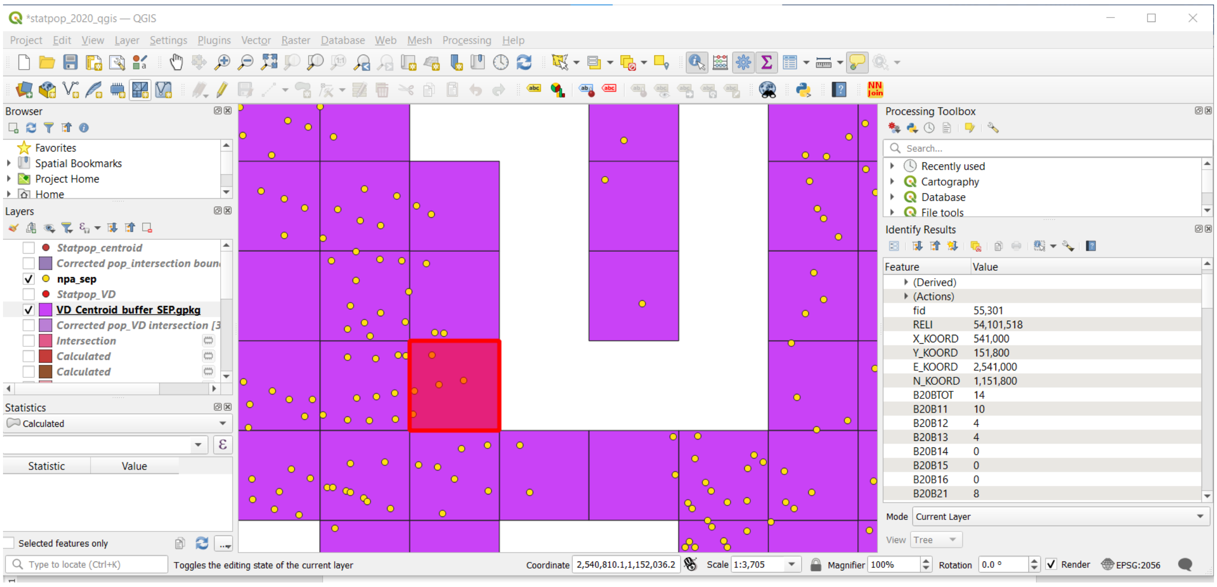

**Supplementary Figure 2B: Flow chart of geocoding procedure of the residential addresses of notifications**

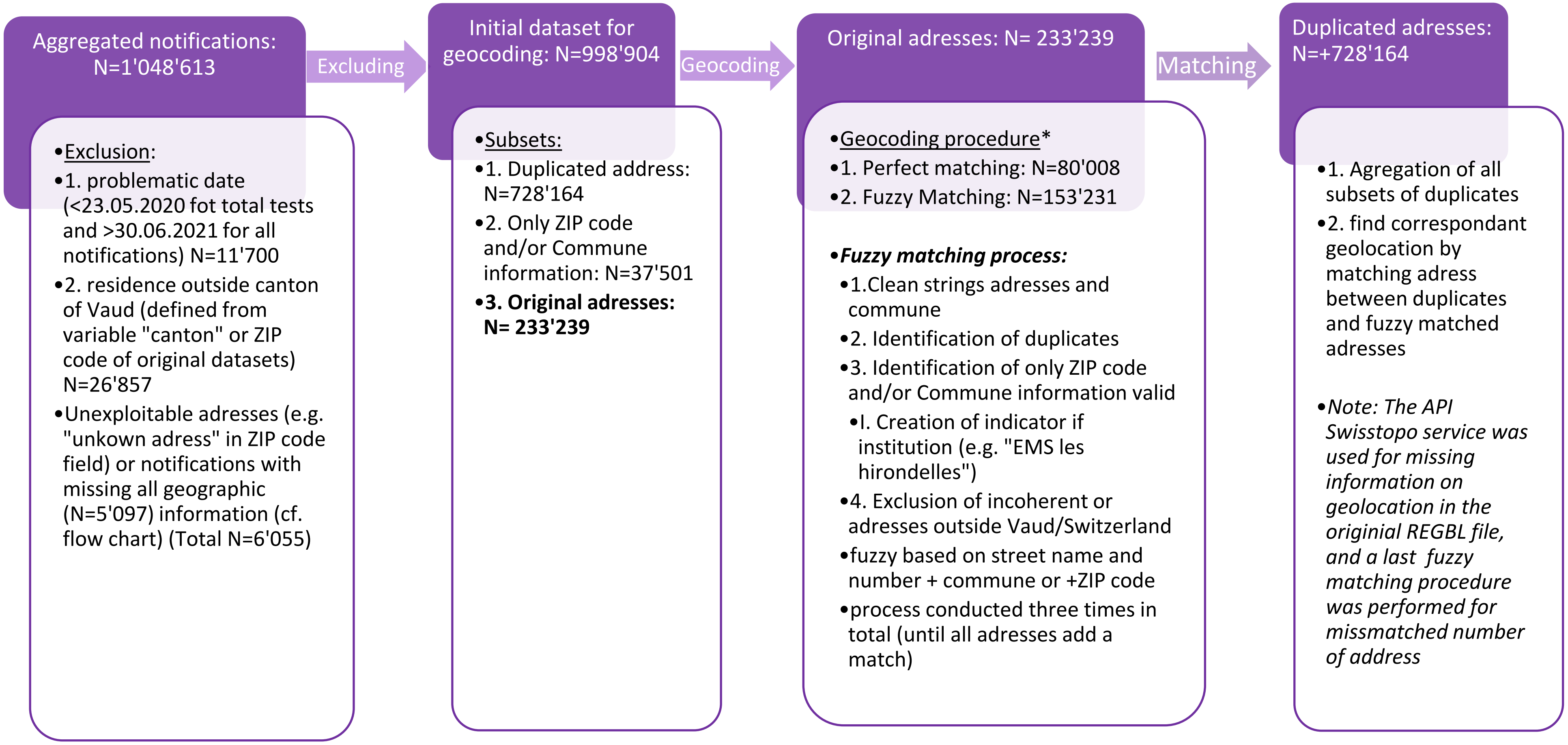

Finally, to establish the necessary dataset for denominator population, we analyzed the distribution of SEP quintiles across age and sex/gender categories within the hectare-level aggregated data and then applied this distribution to the Vaud population dataset, which was disaggregated by age groups and sex categories. This data was obtained from the *Statistique Vaud* (6), specifically the “Permanent resident population by exact age, gender, and origin, Vaud” dataset , as of December 31st, 2020. This dataset, derived from cantonal registers of population, is notably more precise than the GEOSTAT Statpop data. However, in this latter dataset, the cantonal boundaries do not align with the hectometric grid, and the adjustment of population size by proportional coverage within the cantonal boundaries is less than ideal. Additionally, for confidentiality reasons, the GEOSTAT dataset employs a uniform censoring approach for population subgroups with fewer than three individuals, assigning them a standardized value of three. While this practice aims to protect anonymity, it also leads to overestimations in population size.

**Supplementary Section 3: the Swiss socio-economic position (Swiss-SEP)**

The Swiss-SEP is an area-based socio-economic position index centered on residential buildings, encompassing overlapping boundaries. The Swiss-SEP considers neighborhood information based on an average of 50 households (7) and is widely used in social and health research to investigate the associations between social and economic position and health outcomes, as seen in recent studies (8-10). The Swiss-SEP is constructed using principal component analysis, deriving information from the 2000 population census on four indicators as proxies for SEP: median rent per square meter (income proxy), proportion of households led by individuals with a primary education or less (education proxy), proportion of households with individuals in manual or unskilled jobs (occupation proxy), and the average number of persons per room (crowding proxy). The index scores range from 0 to 100, with higher values indicating higher SEP.The Swiss-SEP scores range from 1 to 100, with higher scores indicating a higher SEP. In our study, we utilized the latest updated hybrid version, Swiss-SEP 3 (11), which incorporates information from the 2012 to 2015 annual micro-censuses and updated values for neighborhood centered on buildings constructed after 2000.

Within the geographical boundaries of Vaud, we identified a total of 115’596 SEP neighborhoods. We then matched the residential coordinates of each notification with the nearest SEP neighborhood. SEP index scores were subsequently aggregated into five categories using quintiles as cutoff points, ranging from one (lowest) to five (highest). In cases involving non-residential addresses, such as those corresponding to schools or nursing homes, as well as incomplete addresses, we used the mean SEP of the respective ZIP code area as a proxy for SEP. However, it's noteworthy that notifications related to hospitalization stays and ICU admissions lacked SEP attribution, as these notifications only contained ZIP code information. The reliance on mean SEP from ZIP code areas significantly influenced the distribution within quintiles, leading to a skewed categorization in the lowest and highest quintiles. Therefore, SEP information for hospitalizations and ICU admission outcomes was excluded from our analyses. Given that a significant proportion of death notifications could not be accurately geocoded, analyses using SEP quintiles were likewise restricted to geocoded notifications.

**Supplementary Table 4: Geocoding status of notifications, by COVID-19 outcome**

| | **Tests** **N (%)** N= 885'925 | **Positive tests** **N (%)** N=96'963 | **Hospitalisations** **N (%)** N=6'356 | **ICU admissions** **N (%)** N=1'134 | **Deaths** **N (%)** N=1'175 |
|---|---|---|---|---|---|
| Geocoded | 834'912 (94.2%) | 89'183 (91.9%) | 0 (0%) | 0 (0%) | 713 (60.7%) |
| Incomplete address | 51'013 (4.6%) | 7'780 (6.9%) | 6'359 (100%) | 1'134 (100%) | 462 (21.6%) |
| Non-residential location | 10'779 (1.2%) | 1'157 (1.2%) | NA | NA | 207 (17.7%) |

For both total tests and positive tests, approximately 4.6% and 8%, respectively, could not be geocoded and were therefore attributed to the mean socioeconomic position (SEP) of the respective ZIP code area. Moreover, approximately 1.2% of these notifications corresponded to non-residential addresses (i.e., institutions), receiving a similar SEP attribution based on the ZIP code area.

Regarding deaths, 60.7% of notifications included a complete residential address and were successfully geocoded. Women had a higher proportion of geocodes corresponding to institutional locations (22%), predominantly nursing homes, compared to men (14%).

**Supplementary Figure 5: Flow chart of included notifications**

**Total number of notifications received**

| | Total tests | Positive tests | Hospitalisations | ICU admissions | Deaths |
|---|---|---|---|---|---|
| Total | 938'358 | 100'300 | 7'345 | 1'343 | 1'246 |
| Date range | 02.03.2020-30.06.2021 | 02.03.2020-30.06.2021 | 08.01.2020-30.06.2021 | 04.03.2020-30.06.2021 | 05.03.2020-17.06.2021 |

→

**Ineligible or problematic**

| | Total tests | Positive tests | Hospitalisations | ICU admissions | Deaths |
|---|---|---|---|---|---|
| Problematic date | 20'763[a,c] | 54[c] | 31[b,c] | 1[c] | 0 |
| Residence outside canton of Vaud | 24'339 | 2'071 | 352 | 91 | 4 |

[a] before May 23th 2020
[b] before March 1st 2020
[c] After June 27st 2021

↓

**Eligible**

| Total tests | Positive tests | Hospitalisations | ICU admissions | Deaths |
|---|---|---|---|---|
| 893'256 | 98'175 | 6'962 | 1'251 | 1'242 |

→

**Missing**

| | Total tests N=893'256 | Positive tests N=98'175 | Hospitalisations N=6962 | ICU admissions N=1251 | Deaths N=1242 |
|---|---|---|---|---|---|
| Missing sex | 104 | 63 | 0 | 0 | 1 |
| Sex category "*Other*" | 1'041 | 0 | 0 | 0 | 0 |
| Missing age | 173 | 26 | 2 | 0 | 2 |
| Missing address | 20'814 | 4'733 | 6'966 | 1'252 | 117 |
| Missing commune | 6'993 | 1'107 | 6'966 | 1'252 | 170 |
| Missing zip code | 7'031 | 1'107 | 15 | 3 | 170 |
| Missing all geo data | 5'076(0.6%) | 778 (0.8%) | 15 (0.2%) | 3 (0.2%) | 49 (3.9%) |
| Invalid geo data (defined from geocoding) | 4'110 | 350 | 590 | 114 | 9 |

↓

**Included**

| Total tests | Positive tests | Hospitalisations | ICU admissions | Deaths |
|---|---|---|---|---|
| 885'925 | 96'963 | 6'356 | 1'134 | 1'175 |

**Supplementary Table 6: Incidence rate ratios of total tests, positive tests, hospitalisations, ICU admissions and deaths, negative binomial regression models without interaction terms**

| | *Outcomes* | | | | | |
|---|---|---|---|---|---|---|
| | **Total tests per population** | **Positive tests per population** | **Positive tests per test** | **Hospitalisations per population** | **ICU admissions per population** | **Deaths per population** |
| | IRR (95% CI) | IRR (95% CI) | IRR (95% CI) | IRR (95% CI) | IRR (95% CI) | IRR (95% CI) |
| **(*ref. Men*)** | | | | | | |
| Women | 1.010 (0.98-1.041) | 0.989 (0.953-1.026) | 0.970* (0.945-0.996) | 0.692* (0.488-0.981) | 0.399*** (0.235-0.678) | 0.510*** (0.427-0.608) |
| **(*ref. Age 50-59*)** | | | | | | |
| Age 0-9 | 0.250***(0.235-0.267) | 0.110***(0.101-0.121) | 0.439***(0.408-0.473) | | | |
| Age 10-19 | 0.977 (0.917-1.041) | 0.776***(0.719-0.837) | 0.796***(0.755-0.838) | | | |
| Age 20-29 | 1.438***(1.349-1.532) | 1.185***(1.1-1.277) | 0.823***(0.783-0.866) | | | |
| Age 30-39 | 1.410***(1.323-1.502) | 1.059 (0.983-1.141) | 0.752***(0.714-0.791) | | | |
| Age 40-49 | 1.131***(1.061-1.205) | 1.010 (0.937-1.088) | 0.893***(0.848-0.939) | (ref. Age 0-59) | (ref. Age 0-59) | (ref. Age 0-59) |
| Age 60-69 | 0.881***(0.827-0.939) | 0.861***(0.798-0.929) | 0.976 (0.925-1.028) | 4.833*** (2.855-8.786) | 8.589*** (3.999-21.478) | 29.818*** (15.785-61.282) |
| Age 70-79 | 0.755***(0.709-0.805) | 0.764***(0.707-0.825) | 1.007 (0.954-1.064) | 8.103*** (4.791-14.72) | 11.594*** (5.4-28.986) | 154.327*** (87.466-303.293) |
| Age 80+ | 1.111** (1.042-1.185) | 1.318***(1.22-1.424) | 1.185***(1.122-1.251) | 18.377*** (10.883-33.345) | 9.612*** (4.439-24.171) | 843.06*** (485.617-1638.875) |
| **(ref. Q1) - lowest** | | | | | | |
| 2nd SEP quintile | 1.093*** (1.042-1.147) | 1.033 (0.975-1.095) | 0.942** (0.904-0.981) | NA | NA | 0.954 (0.725-1.254) |
| 3rd SEP quintile | 1.166*** (1.111-1.223) | 1.057 (0.998-1.121) | 0.904*** (0.868-0.942) | NA | NA | 0.982 (0.749-1.289) |
| 4th SEP quintile | 1.241*** (1.183-1.301) | 0.970 (0.915-1.029) | 0.775*** (0.744-0.808) | NA | NA | 0.816 (0.617-1.08) |
| 5th SEP quintile - highest | 1.247*** (1.189-1.308) | 0.897*** (0.846-0.951) | 0.712*** (0.683-0.742) | NA | NA | 0.714* (0.539-0.946) |

Note: SEP= Socioeconomic position; NA= Not Applicable; p-value levels: $^{*}$p<0.05; $^{**}$p<0.01; $^{***}$p<0.001
Regression models: glm.nb(n_*outcome*~SEX+AGE+SEP+offset(log(population))

**Supplementary Table 7: Sensitivity analysis - incidence rate ratios of death notifications using the general population as denominator, employing negative binomial regression models without interaction terms**

As a significant proportion of death notifications could not be accurately geocoded due to incomplete addresses, which either contained only ZIP code information or corresponded to institutional locations, we conducted a sensitivity analysis by including notifications with imprecise SEP attribution and relying on the mean SEP of the ZIP code area.

| | **(1)** | **(2)** | **(3)** |
|---|---|---|---|
| | **Geocoded death notifications, per population** ( N=713) | **Death notifications outside institutions, per population** (N=968) | **All deaths notifications, per population** (N=1175) |
| ***Variables*** | IRR (95% CI) | IRR (95% CI) | IRR (95% CI) |
| **(*ref. Men*)** | | | |
| Women | 0.51*** (0.427-0.608) | 0.514*** (0.439-0.601) | 0.569*** (0.499-0.648) |
| **(*ref. Age 50-59*)** | | | |
| Age 60-69 | 29.818*** (15.785-61.282) | 26.49*** (15.65-47.215) | 25.813*** (15.664-44.534) |
| Age 70-79 | 154.327*** (87.466-303.293) | 124.924*** (78.104-213.853) | 129.063*** (82.797-213.874) |
| Age 80+ | 843.06*** (485.617-1638.875) | 754.753*** (479.575-1276.94) | 824.332*** (536.944-1350.711) |
| ***(ref. Q1)*** | | | |
| 2nd SEP quintile | 0.954 (0.725-1.254) | 1.298* (1.02-1.656) | 1.426** (1.153-1.766) |
| 3rd SEP quintile | 0.982 (0.749-1.289) | 1.218 (0.955-1.557) | 1.643*** (1.335-2.027) |
| 4th SEP quintile | 0.816 (0.617-1.08) | 0.93 (0.721-1.2) | 1.061 (0.849-1.327) |
| 5th SEP quintile - highest | 0.714* (0.539-0.946) | 0.711* (0.547-0.925) | 0.762* (0.603-0.962) |

Note: Death notifications excluding those with incomplete residential addresses (column 1); Excluding institution-related addresses (column 2); Total death notifications (column 3); p-value levels: *p<0.05; **p<0.01; ***p<0.001

Excluding notifications with less precise SEP attribution had an impact on the regression results. When all notifications were included (column 3), our analysis suggested that individuals in the second and third quintiles had higher mortality rates compared to individual living in the lowest SEP areas. We currently lack a definitive explanation for these findings. Conversely, when restricting the analysis to notifications that could be accurately geocoded (column 1), with more precise SEP attribution, these associations became statistically non-significant. Hence, we opted to focus our analysis on death notifications with geocoded residential addresses for accurate SEP attribution.

However, the association between sex/gender and death in the highest quintile remained consistent across all analyses. Individuals in the highest SEP quintile were significantly less likely to die compared to those in the lowest quintile.

**Supplementary Figure 8: Incidence rate ratios of death notifications, stratified for age groups 70-79 and 80+, using Poisson regression models**

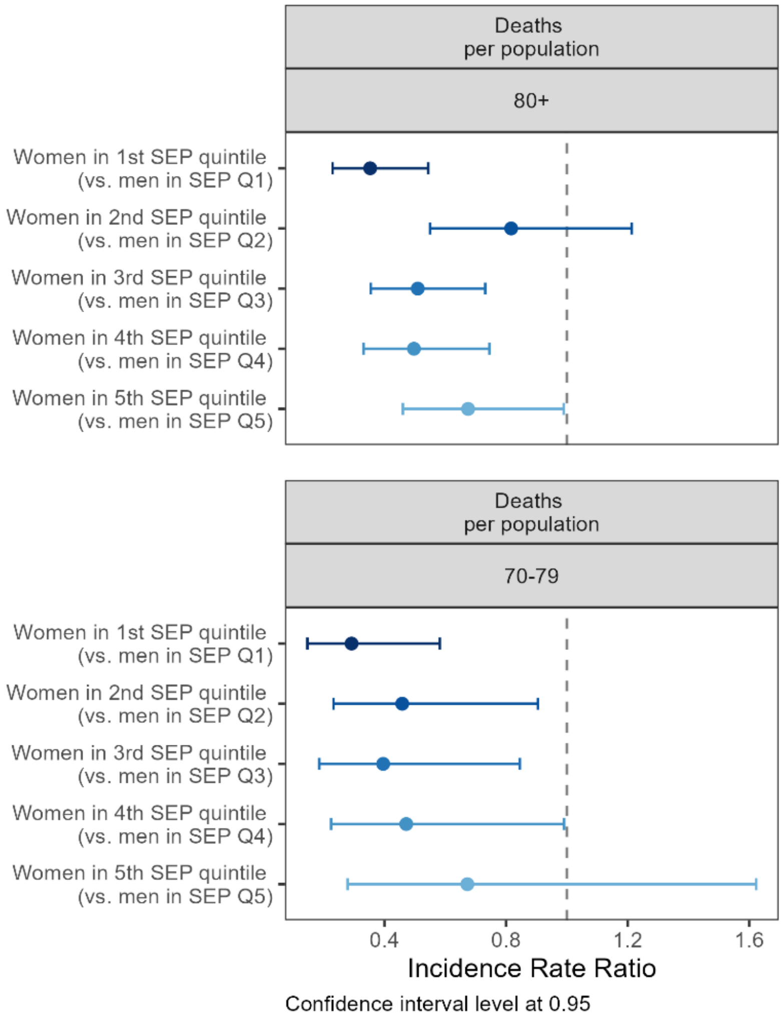


Notes: This figure illustrates incidence rate ratios of administrative sex (ref.: men) for deaths, stratified by quintiles of socio-economic position (SEP), for age groups 80+ (upper part), and 70-79 (lower part). Results for age groups 0-59 and 60-69 are not presented due to the absence of significant differences, attributed to a small number of events in these categories. Estimation was obtained using a Poisson regression model as no overdispersion was found in this triple interaction model.

The relationships between sex/gender, age, and SEP were investigated using a triple interaction term across these covariates. Findings revealed that, globally, for both age groups 70-79 and 80+, the sex/gender gap in mortality decreased with increasing SEP.

**Supplementary Figure 9: Sensitivity analysis - incidence rate ratios of death notifications using the general population as denominator, employing negative binomial regression models with interaction terms**

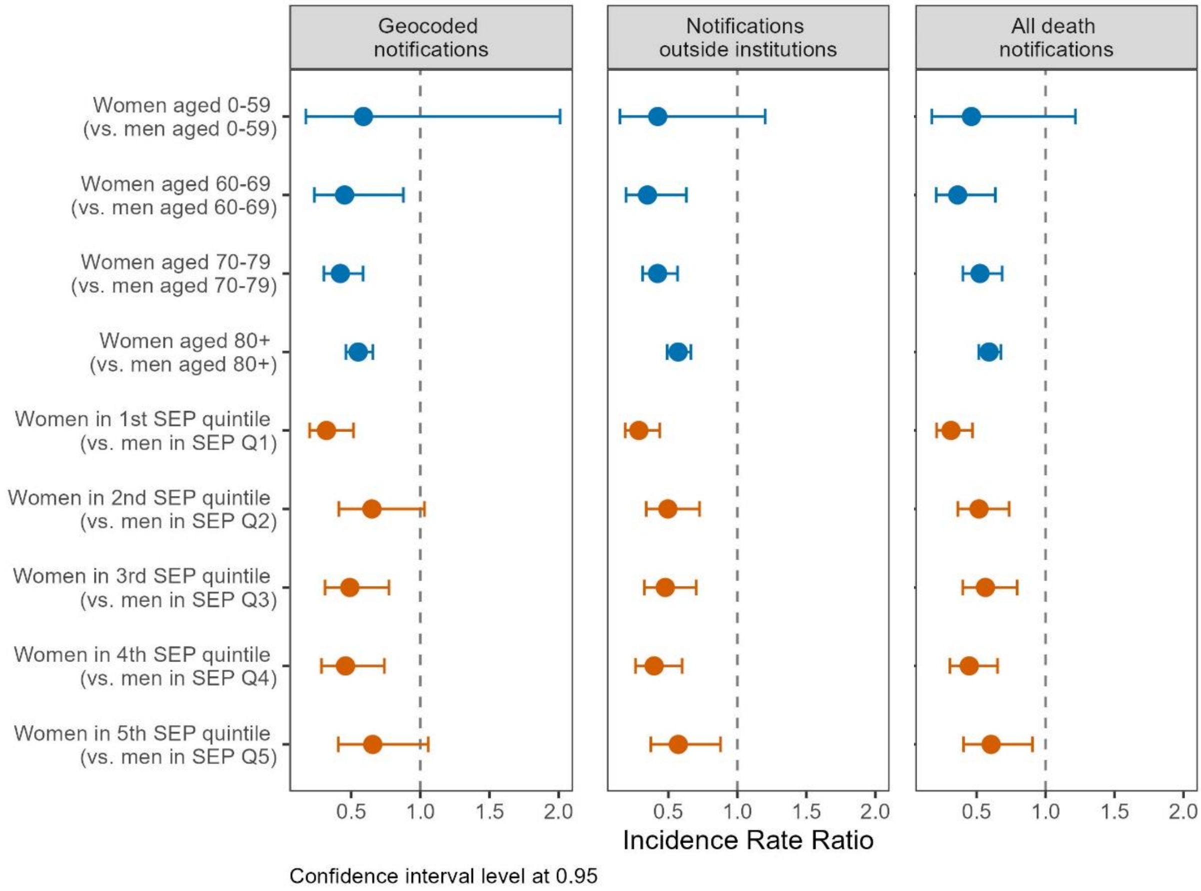


**Figure S9 – Incidence rate ratios of administrative sex (ref.: men) for geocoded death notifications (column 1), notifications of death including institutional location (column 2) and all death notifications (column 3), stratified by age groups (upper part), and quintiles of socio-economic position (SEP, lower part), using general population as denominator, Canton of Vaud surveillance data 2020-2021, Switzerland**

Notes: Geocoded death notifications (column 1), N=713 N; including institution-related addresses (column 2), N=968; including incomplete addresses without accurate retrievable SEP (column 3), =1175.

Coefficients of negative binomial models using interaction term between sex/gender and age and/or SEP quintiles reported in forest plots were estimated using the *contrast* function of the *emmeans*, for *estimated marginal means*, package in R (12). This function facilitates the pairwise comparisons by treating each category of age and SEP as the reference category, comparing women to men.

### Reference

1. Office Fédéral de la Statistique. Registre fédéral des bâtiments et des logements (RegBL): Office Fédéral de la Statistique (2022) [Available from: https://www.housing-stat.ch/fr/madd/public.html.
2. Robinson D, Elias J. Package 'fuzzyjoin'. URL: https://cran r-project org/web/packages/fuzzyjoin/fuzzyjoin pdf. (2018).
3. Pebesma EJ. Simple features for R: standardized support for spatial vector data. R J. (2018);10(1):439.
4. Population and Household Statistics Geodata (STATPOP GEOSTAT) (2020). Federal Office of Statistics; [Available from: https://www.bfs.admin.ch/bfs/en/home/services/geostat/swiss-federal-statistics-geodata/population-buildings-dwellings-persons/population-housholds-from-2010.html.
5. QGIS Development Team. QGIS Geographic Information System. Open Source Geospatial Foundation Project; (2022).
6. Statistique Vaud (2020). Population résidante permanente par âge exact_ sexe et origine_ Vaud_ 2017-2020 [Available from: https://www.vd.ch/themes/etat-droit-finances/statistique/statistiques-par-domaine/01-population/etat-et-structure-de-la-population.
7. Panczak R, Galobardes B, Voorpostel M, Spoerri A, Zwahlen M, Egger M. A Swiss neighbourhood index of socioeconomic position: development and association with mortality. J Epidemiol Community Health. (2012);66(12):1129-36.
8. Ihle A, Gabriel R, Oris M, Gouveia ÉR, Gouveia BR, Marques A, et al. Cognitive reserve mediates the relation between neighborhood socio-economic position and cognitive decline. Dementia and Geriatric Cognitive Disorders Extra. (2022);12(2):90-3.
9. Matthes KL, Zuberbuehler CA, Rohrmann S, Hartmann C, Siegrist M, Burnier M, et al. Selling, buying and eating–a synthesis study on dietary patterns across language regions in Switzerland. British Food Journal. (2021);124(5):1502-18.
10. Jeong A, Eze IC, Vienneau D, de Hoogh K, Keidel D, Rothe T, et al. Residential greenness-related DNA methylation changes. Environ Int. (2022);158:106945.
11. Panczak R, Berlin C, Voorpostel M, Zwahlen M, Egger M. The Swiss neighbourhood index of socioeconomic position: update and re-validation. Swiss Medical Weekly. (2023);153(1):40028.
12. Lenth RV. Emmeans: Estimated Marginal Means, aka Least-Squares Means (2023) [Available from: https://CRAN.R-project.org/package=emmeans.
